# Leveraging Northern European population history; novel low frequency variants for polycystic ovary syndrome

**DOI:** 10.1101/2021.05.20.21257510

**Authors:** Jaakko S. Tyrmi, Riikka K. Arffman, Natàlia Pujol-Gualdo, Venla Kurra, Laure Morin-Papunen, Eeva Sliz, FinnGen, Estonian Biobank Research Team, Terhi T. Piltonen, Triin Laisk, Johannes Kettunen, Hannele Laivuori

## Abstract

**Background:** Polycystic ovary syndrome (PCOS) is a common, complex disorder, which should be recognized as a prominent health concern also outside the context of fertility. Although PCOS affects up to 18% of women worldwide, its etiology remains poorly understood. It is likely that a combination of genetic and environmental factors contributes to the risk of PCOS development. Whilst previous genome-wide association studies have mapped several loci associated with PCOS, analysis of populations with unique population history and genetic makeup has the potential to uncover new low frequency variants with larger effects. In this study, we leverage genetic information of two neighboring and well-characterized populations in Europe – Finnish and Estonian – to provide a basis for a new understanding of the genetic determinants of PCOS.

**Methods and Findings:** We conducted a three-stage case-control genome-wide association study (GWAS). In the discovery phase, we performed a GWAS comprising of a total of 797 cases and 140,558 controls from the FinnGen study. For validation, we used an independent dataset from the Estonian Biobank, including 2,812 cases and 89,230 controls. Finally, we conducted a joint meta-analysis of 3,609 cases and 229,788 controls from both cohorts.

In total, we identified three novel genome-wide significant variants associating with PCOS. Two of these novel variants, rs145598156 (p=3.6 × 10^−8^, OR=3.01 [2.02-4.50] MAF=0.005) and rs182075939 (p=1.9 × 10^−16^, OR= 1.69 [1.49-1.91], MAF=0.04), were found to be enriched in the Finnish and Estonian populations and are tightly linked to a deletion c.1100delC (r^2^= 0.95) and a missense I157T (r^2^=0.83) in *CHEK2*. The third novel association is a common variant near *MYO10* (rs9312937, p= 1.7 × 10^−8^, OR=1.16 (1.10-1.23), MAF=0.44). We also replicated four previous reported associations near the genes *ERBB4, DENND1A, FSHB* and *ZBTB16*.

**Conclusions:** We identified three novel variants for PCOS in a Finnish-Estonian GWAS. Using isolated populations to perform genetic association studies provides a useful resource to identify rare variants contributing to the genetic landscape of complex diseases such as PCOS.

## Introduction

Polycystic ovary syndrome (PCOS) is a common, multifaceted endocrine disorder with a high risk of comorbidity. The recently published international evidence-based guideline recommends using the Rotterdam criteria for PCOS diagnosis. This requires the presence of at least two of the three following symptoms for diagnosis: oligo- or anovulation, clinical or biochemical hyperandrogenism, or polycystic ovaries seen in ultrasound, after exclusion of related disorders [1] This criterion results into a prevalence as high as 18% for PCOS among fertile aged women [2,3], and produces several phenotypes.

PCOS is the most common cause for anovulatory infertility caused by disrupted follicle development due to dysregulation in the hypothalamus-pituitary-axis. This results in follicle arrest and an increase in the number of antral follicles in the ovaries, as well as a 2-3-fold increase in levels of anti-müllerian hormone (AMH). [4] Ovulatory dysfunction often subsides with age; however, women with PCOS still display higher AMH and a later onset of menopause [5-9]. In addition to the reproductive features, PCOS is also characterized by metabolic disturbances such as obesity, insulin resistance, and dyslipidemia [10-12]. Women with PCOS also have an increased risk for endometrial cancer; however, the majority of studies do not indicate a higher susceptibility to other types of cancer [13-16].

Despite the high prevalence of the syndrome, the origins of PCOS remain unknown. Considering the complex nature of the syndrome, it is likely that both genetic and environmental factors contribute to the risk for its development. Non-genetic factors for PCOS include prenatal androgen exposure, early weight gain, insulin resistance, and low levels of sex hormone binding globulin (SHBG). [17-19]

Notably, the heritability of PCOS is estimated to be around 70% [20,21]. To elucidate the genetic architecture of PCOS, several genome-wide association studies (GWAS) and meta-analysis studies have been conducted and have mapped over 20 susceptibility loci for PCOS [22-30]. The identified loci indicate roles in PCOS for gonadotrophin signaling, folliculogenesis, epithelial growth factor signaling, DNA repair and structure, cell cycle and proliferation, and androgen biosynthesis. However, these common genetic variants explain only around 10% of the heritability [31]. Thus, it has been suggested that rare variants with larger effect sizes may contribute to the heritability of PCOS [32]. Nevertheless, the identification of these may be difficult in data sets with large genetic variation.

The value of studying genetic isolates, such as the Finnish population, has been understood early on before the era of genome-wide association studies [33]. Such populations provide an excellent opportunity to facilitate the discovery of rare variants with larger effects and characterize the genetic basis of complex diseases such as PCOS. The Finnish population originates from a small founder population who underwent several bottleneck events occurring over centuries, followed by genetic drift. These events have led to the enrichment of many low frequency variants, including loss-of-function mutations and gene knockouts, that are almost absent in many other European populations [34-36]. Replication of association results may be difficult when studying isolated populations as, by definition, closely related populations are uncommon. In the case of the Finns, the Estonian population provides a natural comparison, as it is genetically the closest population [34,37].

In this study, we first utilized genome-wide association analyses and the data available in FinnGen project and the Estonian Biobank (EstBB) to unravel novel variants in these population isolates that might provide new insight into the origin and pathophysiology of PCOS. As hyperandrogenism is considered one of the hallmarks of PCOS, and SHBG controls the bioavailability of sex hormones, we then separately searched for significant associations with PCOS in a set of variants that had been recently associated with high total testosterone, bioavailable testosterone, or SHGB levels in women [38]. Finally, as several studies suggest a causal role for obesity in PCOS [39-41], we examined the influence of body mass index (BMI) to our reported associations with PCOS.

As a result, we unraveled two rare and population enriched variants located in the Checkpoint kinase 2 (*CHEK2*) gene and described one novel variant in the intron of the myosin X (*MYO10*) gene. Additionally, we replicated previously reported associations for Erb-B2 Receptor Tyrosine Kinase 4 (ERBB4), DENN Domain Containing 1A (*DENND1A*), Follicle Stimulating Hormone Subunit Beta (*FSHB*) and Zinc Finger And BTB Domain Containing 16 (*ZBTB16*). Moreover, by utilizing a set of variants previously reported to be significantly associated with testosterone levels in women, we detected an additional novel association with PCOS in an intron of Pleckstrin Homology Domain Containing M3 (*PLEKHM3*).

## Materials and Methods

This study is reported according to the Strengthening the Reporting of Genetic Association Studies (STREGA) guideline (Supplementary Checklist 1).

### Study cohorts

#### FinnGen

The FinnGen study combines genotype data from the Finnish biobanks with the digital health record data from the Care Register for Health Care (from 1968 onwards) and the cancer (1953-), cause of death (1969-), and medication reimbursement (1995-) registries (https://www.finngen.fi/en). FinnGen data freeze release 6 (R6) combines the genomic information of 141,355 women (6% of the female Finnish population). In FinnGen, cases of PCOS were defined as women with a record of the following International Classification of Diseases (ICD)-10 code E28.2, ICD-9 code 256.4, or ICD-8 code 256.90. Controls were all women without a PCOS diagnosis, and no other exclusions were made. With this definition, there were 797 cases and 140,558 controls.

Patients and control subjects in FinnGen provided informed consent for biobank research, based on the Finnish Biobank Act. Alternatively, older research cohorts, collected prior the start of FinnGen (in August 2017), were collected based on study-specific consents and later transferred to the Finnish biobanks after approval by the National Supervisory Authority for Welfare and Health, Fimea. Recruitment procedures followed the biobank protocols approved by Fimea. The Coordinating Ethics Committee of the Hospital District of Helsinki and Uusimaa (HUS) approved the FinnGen study protocol (Nr HUS/990/2017).

The FinnGen study was approved by Finnish Institute for Health and Welfare (permit numbers: THL/2031/6.02.00/2017, THL/1101/5.05.00/2017, THL/341/6.02.00/2018, THL/2222/6.02.00/2018, THL/283/6.02.00/2019, THL/1721/5.05.00/2019, THL/1524/5.05.00/2020, and THL/2364/14.02/2020); Digital and population data service agency (permit numbers: VRK43431/2017-3, VRK/6909/2018-3, VRK/4415/2019-3); the Social Insurance Institution (permit numbers: KELA 58/522/2017, KELA 131/522/2018, KELA 70/522/2019, KELA 98/522/2019, KELA 138/522/2019, KELA 2/522/2020, KELA 16/522/2020); and Statistics Finland (permit numbers: TK-53-1041-17 and TK-53-90-20).

The Biobank access decisions for FinnGen samples and data utilized in the FinnGen Data Freeze 6 include: THL Biobank BB2017_55, BB2017_111, BB2018_19, BB_2018_34, BB_2018_67, BB2018_71, BB2019_7, BB2019_8, BB2019_26, BB2020_1, Finnish Red Cross Blood Service Biobank 7.12.2017, Helsinki Biobank HUS/359/2017, Auria Biobank AB17-5154, Biobank Borealis of Northern Finland_2017_1013, Biobank of Eastern Finland 1186/2018, Finnish Clinical Biobank Tampere MH0004, Central Finland Biobank 1-2017, and Terveystalo Biobank STB 2018001.

#### Estonian Biobank

The Estonian Biobank (EstBB) is a population-based biobank with over 200,000 participants, currently including approximately 135,000 women (20% of the female Estonian population). The 150K data freeze was used for the analyses described in this paper. All biobank participants have signed a broad informed consent form. Individuals with PCOS were identified using the ICD-10 code E28.2, and all female biobank participants who did not have this diagnosis served as controls. This included a total of 2,812 cases and 89,230 controls. Information on the ICD codes was obtained via regular linking with the national Health Insurance Fund and other relevant databases [42]. Analyses in the EstBB were carried out under ethical approval 1.1-12/624 from the Estonian Committee on Bioethics and Human Research and data release N05 from the EstBB.

### Genotyping and association analyses

#### FinnGen

Sample genotyping in FinnGen was performed using Illumina and Affymetrix arrays (Illumina Inc., San Diego, and Thermo Fisher Scientific, Santa Clara, CA, USA). Genotype calls were made using GenCall or zCall [43] for Illumina and the AxiomGT1 algorithm for Affymetrix data. Genotypes with a Hardy-Weinberg Equilibrium (HWE) p-value below 1e-6, minor allele count < 3, and genotyping success rate < 98 % were removed. Samples with ambiguous gender, those with high genotype missingness > 5%, and those that were outliers in the population structure (> 4 SD from the mean on first two dimensions of PCA) were omitted. Samples were pre-phased with Eagle 2.3.5 [44] using 20,000 conditioning haplotypes. Genotypes were imputed with Beagle 4.1 [45] using the SiSu v3 imputation reference panel, which consisted of 3,775 individuals of Finnish ancestry with sequenced whole genomes. The post-imputation protocol is publicly available at https://dx.doi.org/10.17504/protocols.io.xbgfijw.

Association analysis was performed using a generalized mixed model as implemented in SAIGE [46]. Included adjustments were age, genotyping batches, and the first ten principal components (PCs).

Formatting and preparation of the FinnGen association data for downstream analysis were managed with worfkflow management software STAPLER [47].

#### Estonian Biobank

All EstBB participants were genotyped using Illumina GSAv1.0, GSAv2.0, and GSAv2.0_EST arrays at the Core Genotyping Lab of the Institute of Genomics, University of Tartu. Samples were genotyped and PLINK format files were created using Illumina GenomeStudio v2.0.4. Individuals were excluded from the analysis if their call-rate was < 95% or if their sex defined by heterozygosity of X chromosomes did not match their sex in the phenotype data. Before imputation, variants were filtered by call-rate < 95%, HWE p-value < 1e-4 (autosomal variants only), and minor allele frequency < 1%. Variant positions were updated to b37 and all variants were changed to be from the TOP strand using GSAMD-24v1-0_20011747_A1-b37.strand.RefAlt.zip files from the https://www.well.ox.ac.uk/~wrayner/strand/ webpage. Pre-phasing was conducted using Eagle v2.3 software [44] (number of conditioning haplotypes Eagle2 uses when phasing each sample was set to: --Kpbwt=20000) and imputation was done using Beagle v.28Sep18.793 [45] with effective population size ne=20,000. The population specific imputation reference of 2,297 whole genome sequencing (WGS) samples was used [48].

Association analysis was carried out using SAIGE (v0.38) software to implement a mixed logistic regression model with year of birth and 10 PCs as covariates in step I. A total of 2,812 cases and 89,230 controls were included in the analyses.

### Meta-analysis

In order to synchronize the build of the datasets, we lifted the FinnGen GWAS summary statistics over to hg37 build using UCSC liftOver [49] before running the meta-analyses. METAL software was used to perform inverse variance-weighted meta-analysis for FinnGen and EstBB GWAS results [50]. In total, 3,609 cases and 229,788 controls were analyzed. High imputation quality markers (INFO score > 0.7) were kept from each study prior to the meta-analysis. A total of 24,157,216 markers were included in the analysis. Genome-wide significance was set to p < 5 × 10^−8^. The meta-analyses were conducted independently by two analysts, and summary statistics were compared for consistency.

### Functional annotation and gene prioritization

In order to identify plausible candidate genes we used the FUMA platform [51]. FUMA uses GWAS summary statistics and performs extensive functional annotation and candidate gene mapping using positional, expression Quantitative Trait Loci (eQTL), and chromatin interaction (HiC) mapping in all genome-wide significant loci. Loci were defined by ±1000 kb of the top single nucleotide variant (SNV) in the region. Gene-based analysis was also performed in this platform using MAGMA [52]. We prioritized variants that were more likely to have a functional consequence, such as variants in high linkage disequilibrium (LD) (r2>0.6) with missense mutations or pathogenic variants. Secondly, we prioritized variants overlapping with regulatory marks, focusing on genes with modified expression or genes that showed chromatin interaction links with the variants. Furthermore, gene functions were examined in GenBank and UniProt portals. In addition, a literature search was performed for the genes of interest to gain further insight into the possible underlying molecular mechanisms. Those genes showing relevant functions in relevant tissues or traits with similar PCOS pathophysiology were ultimately considered for gene candidate prioritization.

### Colocalization analyses

We tested whether the GWAS signals colocalized with variants that affect gene expression using the following pipeline (https://github.com/eQTL-Catalogue/colocalisation)[53]. We compared our significant loci to all eQTL Catalogue RNA-Seq datasets containing QTLs for gene expression, exon expression, transcript usage, and txrevise event usage; eQTL Catalogue microarray datasets containing QTLs for gene expression; and GTEx v7 datasets containing QTLs for gene expression [53]. We lifted the GWAS summary statistics over to hg38 build in order to match the eQTL catalogue and convert the summary statistics to VCF format. For each genome-wide significant (p<5 ×10^−8^) GWAS variant, we extracted the 1Mbp radius of its top hit from the QTL datasets. We then ran the colocalization analysis for those eQTL catalogue traits that had at least one cis-QTL within this region with p< 1×10^−6^. We considered two signals to colocalize if the posterior probability for a shared causal variant was 0.8 or higher.

### Conditional analyses

Since considering most significant variants as the causal ones would lead to an underestimation of the total variance explained at each locus, we next performed conditional analyses, which were carried out similarly to the main association testing using SAIGE [46]. This approach has been used to identify secondary association signals at a particular locus and involves association analysis conditioning on the primary associated variant at the locus to test whether there are any additional significantly associated variants [54]. We proceeded to test associations using a step-wise analysis, where markers were added to the model until no independent signals were identified.

### Adjusting the GWAS for BMI

To investigate the influence of body mass index (BMI) on PCOS, we ran an additional association analysis including BMI as a covariate. In the discovery dataset, this analysis contained a total of 482 PCOS cases (60.5 % of the original PCOS sample) and 91,631 controls from FinnGen (65.2 % of the original control sample). Similarly, we ran an association analysis including BMI as a covariate for the validation dataset, which contained a total of 2,137 PCOS cases (75% of the original PCOS sample size) and 68,690 controls from EstBB (76.9 % of the original control sample size). We then performed a second meta-analysis including the two GWAS adjusted for BMI from both cohorts. This analysis included 2,619 cases and 160,321 controls, and a total of 24,461,102 genetic markers were analyzed.

### Interaction analysis

Previous studies have shown that patients with invasive breast cancer who are carriers of the c.1100delC mutation are more likely to be obese, though this is not the case for the general population [53]. Thus, we tested whether a similar interaction can be seen with PCOS. We fitted a logistic model where PCOS was the outcome, the lead variant genotype and BMI formed the interaction term, and the ten first genetic PCs along with age were added as covariates. This analysis was performed with R version 4.0.5.

### Search for variants previously associated with testosterone and SHBG levels

Hyperandrogenism is considered one of the hallmarks of PCOS, and SHBG controls the bioavailability of sex hormones. Thus, we separately searched for significant associations with PCOS in a set of candidate variants that were recently associated with high total testosterone, bioavailable testosterone, or SHGB levels in women [54]. In total 217, 154, and 304 of our meta-analyzed variants overlapped with high total testosterone, bioavailable testosterone, and SHBG level associations, respectively. Statistical significance levels were defined using the Bonferroni correction, with a Bonferroni-adjusted threshold of association defined as 0.05/(217 + 154 + 304) =7.4×10^−5^.

### Data availability

Full meta-analysis summary statistics will be made available upon publication.

## Results

### Discovery GWAS identified a rare novel association for PCOS in *CHEK2*

FinnGen GWAS uncovered two loci, close to *ERBB4* and *DENND1A* that had been previously shown to be associated with PCOS. In addition, a previously unreported large effect association was found in chromosome 22 at 22q11 (Fig 1A).

**Fig 1.**
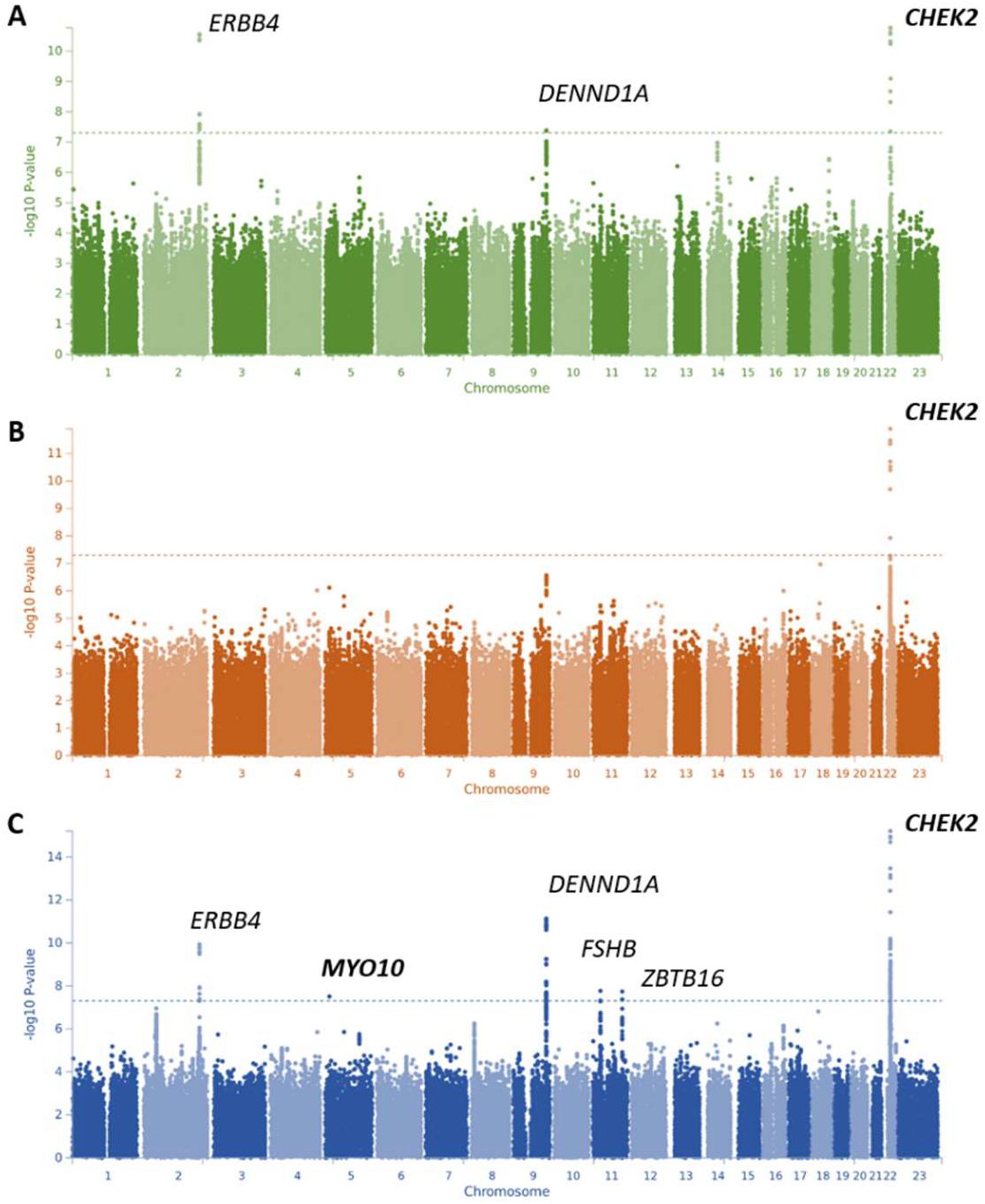
Manhattan plot of the results from the age-adjusted GWAS from the Finnish dataset (A), GWAS from Estonian dataset (B) and joint GWAS meta-analysis of PCOS (C). The novel gene candidates in the six genome-wide significant loci are highlighted in bold. The y axis represents -log(two-sided P values) for association of variants with PCOS from meta-analysis, using an inverse-variance weighted fixed effects model. The horizontal dashed line represents the threshold for genome-wide significance.

The lead variant rs145598156 [p=1.7 × 10^−11^, OR=11.63 (5.69-23.77)] is located in an intronic region 11 kb from the transcription start site (TSS) of ZNFR3 (Table 1, Fig 2A). However, the tight linkage disequilibrium (LD) spans an area of approximately 2Mbp surrounding the lead variant with many variants in high LD (Fig 2A). Functional characterization of this locus revealed a frameshift variant, c.1100delC [rs555607708, p= 1.68 × 10^−9^, OR=13.46 (5.68-31.89)] in CHEK2, with a high LD (r2 = 0.95) with the lead variant. Interestingly, the protein truncating variant c.1100delC is enriched in the Finnish population (AF=0.008) compared to the Estonian (0.003) and other European populations (AF=0.002), according to the gnomAD database [55]. The analysis conditioned on c.1100delC resulted in no genome-wide significant associations in this locus, with a p-value of 3.29 × 10^−4^ for the lead variant rs145598156 (Fig 2B).

**Table 1.** Summary of association results of the genome-wide association meta-analysis of PCOS.

| SNP | Chr:BP | Cytoband | EA/<br>NEA | Nearest<br>gene | Candidate<br>gene | Cohort | EAF<br>(%) | *OR (95% CI) | *P | **OR (95% CI) | **P |
| --- | --- | --- | --- | --- | --- | --- | --- | --- | --- | --- | --- |
| <u>rs873779</u> | <u>chr2:</u><br><u>208773860</u> | <u>2q33</u> | <u>T/C</u> | <u>PLEKHM3</u> | <u>FZD5</u> | FinnGen | 47.90 | 1.10 (1.00-1.22) | 5.7x10<sup>-02</sup> | 1.16 (1.02-1.32) | 2.2x10<sup>-02</sup> |
|  |  |  |  |  |  | EstBB | 53.44 | 1.12 (1.06-1.19) | 1.1x10<sup>-04</sup> | 1.14 (1.07-1.22) | 4.4x10<sup>-04</sup> |
|  |  |  |  |  |  | *Meta* | *52.04* | *1.12 (1.06-1.18)* | *1.7x10<sup>-05</sup>* | *1.15 (1.09-1.21)* | *2.9x10<sup>-06</sup>* |
| rs7564590 | chr2:<br>213387900 | 2q34 | T/C | ERBB4 | ERBB4 | FinnGen | 34.50 | 1.43 (1.29-1.59) | 3.0x10<sup>-11</sup> | 1.50 (1.31-1.72) | 4.4x10<sup>-09</sup> |
|  |  |  |  |  |  | EstBB | 35.87 | 1.12 (1.06-1.19) | 1.1x10<sup>-04</sup> | 1.13 (1.05-1.20) | 2.5x10<sup>-04</sup> |
|  |  |  |  |  |  | *Meta* | *35.56* | *1.19 (1.13-1.25)* | *4.8x10<sup>-11</sup>* | *1.19 (1.13-1.25)* | *4.6x10<sup>-09</sup>* |
| <u>rs9312937</u> | <u>chr5:</u><br><u>16836005</u> | <u>5p15</u> | <u>C/T</u> | <u>MYO10</u> | <u>MYO10</u> | FinnGen | 42.00 | 1.15 (1.04-1.27) | 6.8x10<sup>-03</sup> | 1.06 (0.93-1.20) | 3.7x10<sup>-01</sup> |
|  |  |  |  |  |  | EstBB | 45.47 | 1.16 (1.10-1.23) | 7.7x10<sup>-07</sup> | 1.18 (1.10-1.26) | 1.5x10<sup>-06</sup> |
|  |  |  |  |  |  | *Meta* | *44.58* | *1.16 (1.10-1.22)* | *1.7x10<sup>-08</sup>* | *1.15 (1.08-1.22)* | *3.0x10<sup>-06</sup>* |
| rs3945628 | chr9:<br>126535553 | 9q33 | C/T | DENND1A | DENND1A | FinnGen | 6.64 | 1.74 (1.42-2.15) | 1.5x10<sup>-07</sup> | 1.68 (1.29-2.19) | 9.3x10<sup>-05</sup> |
|  |  |  |  |  |  | EstBB | 7.08 | 1.33 (1.19-1.48) | 2.7x10<sup>-07</sup> | 1.32 (1.17-1.49) | 6.9x10<sup>-06</sup> |
|  |  |  |  |  |  | *Meta* | *6.99* | *1.40 (1.27-1.55)* | *2.9x10<sup>-12</sup>* | *1.38 (1.23-1.54)* | *1.0x10<sup>-08</sup>* |
| rs11031002 | chr11:<br>30215261 | 11p14 | A/T | FSHB | FSHB | FinnGen | 12.00 | 1.33 (1.14-1.56) | 2.3x10<sup>-04</sup> | 1.31 (1.29-2.19) | 5.7x10<sup>-03</sup> |
|  |  |  |  |  |  | EstBB | 12.27 | 1.22 (1.12-1.32) | 5.8x10<sup>-06</sup> | 1.16 (1.05-1.27) | 2.2x10<sup>-03</sup> |
|  |  |  |  |  |  | *Meta* | *12.21* | *1.24 (1.15-1.34)* | *9.2x10<sup>-09</sup>* | *1.18 (1.10-1.27)* | *7.7x10<sup>-05</sup>* |
| rs1672716 | chr11:<br>113952497 | 11q23 | G/A | ZBTB16 | ZBTB16 | FinnGen | 14.60 | 0.74 (0.64-0.86) | 5.2x10<sup>-05</sup> | 0.78 (0.65-0.94) | 1.0x10<sup>-02</sup> |
|  |  |  |  |  |  | EstBB | 14.49 | 0.84 (0.77-0.91) | 1.7x10<sup>-05</sup> | 0.81 (0.74-0.89) | 1.4x10<sup>-05</sup> |
|  |  |  |  |  |  | *Meta* | *14.51* | *0.81 (0.76-0.87)* | *9.8x10<sup>-09</sup>* | *0.80 (0.74-0.87)* | *4.7x10<sup>-07</sup>* |
| <u>rs145598156</u> | <u>chr22:</u><br><u>29416402</u> | <u>22q12</u> | <u>T/C</u> | <u>ZNFR3</u> | <u>CHEK2</u> | FinnGen | 0.79 | 11.63 (5.69-23.77) | 1.7x10<sup>-11</sup> | 13.5 (5.35-34.38) | 4.5x10<sup>-08</sup> |
|  |  |  |  |  |  | EstBB | 0.37 | 1.68 (1.05-2.69) | 3.2x10<sup>-02</sup> | 1.53 (0.90-2.61) | 1.1x10<sup>-01</sup> |
|  |  |  |  |  |  | *Meta* | *0.52* | *3.01 (2.02-4.50)* | *3.6x10<sup>-08</sup>* | *2.61 (2.14-3.07)* | *4.4x10<sup>-05</sup>* |
| <u>rs182075939</u> | <u>chr22:</u><br><u>29098376</u> | <u>22q12</u> | <u>G/A</u> | <u>TTC28</u> | <u>CHEK2</u> | FinnGen | 3.19 | 1.95 (1.44-2.65) | 1.8x10<sup>-05</sup> | 2.18 (1.46-3.24) | 1.1x10<sup>-04</sup> |
|  |  |  |  |  |  | EstBB | 4.64 | 1.64 (1.43-1.88) | 1.3x10<sup>-12</sup> | 1.68 (1.44-1.96) | 4.6x10<sup>-11</sup> |
|  |  |  |  |  |  | *Meta* | *4.41* | *1.69 (1.49-1.91)* | *1.9x10<sup>-16</sup>* | *1.74 (1.5-2.01)* | *4.9x10<sup>-14</sup>* |

*P*, P-value; *OR*, odds ratio; *EAF*, effect allele frequency; *EA*, effect allele; *NEA*, non-effect allele. Meta-analysis results of the GWA-studies from Estonian Biobank (EstBB) and FinnGen are shown in italics. Novel associations are underlined. Variant positions (BP) are according to GRCh37/hg19.
\* OR and P-values of age adjusted results
\*\* OR and P-values of age and BMI adjusted results

**Fig 2.**
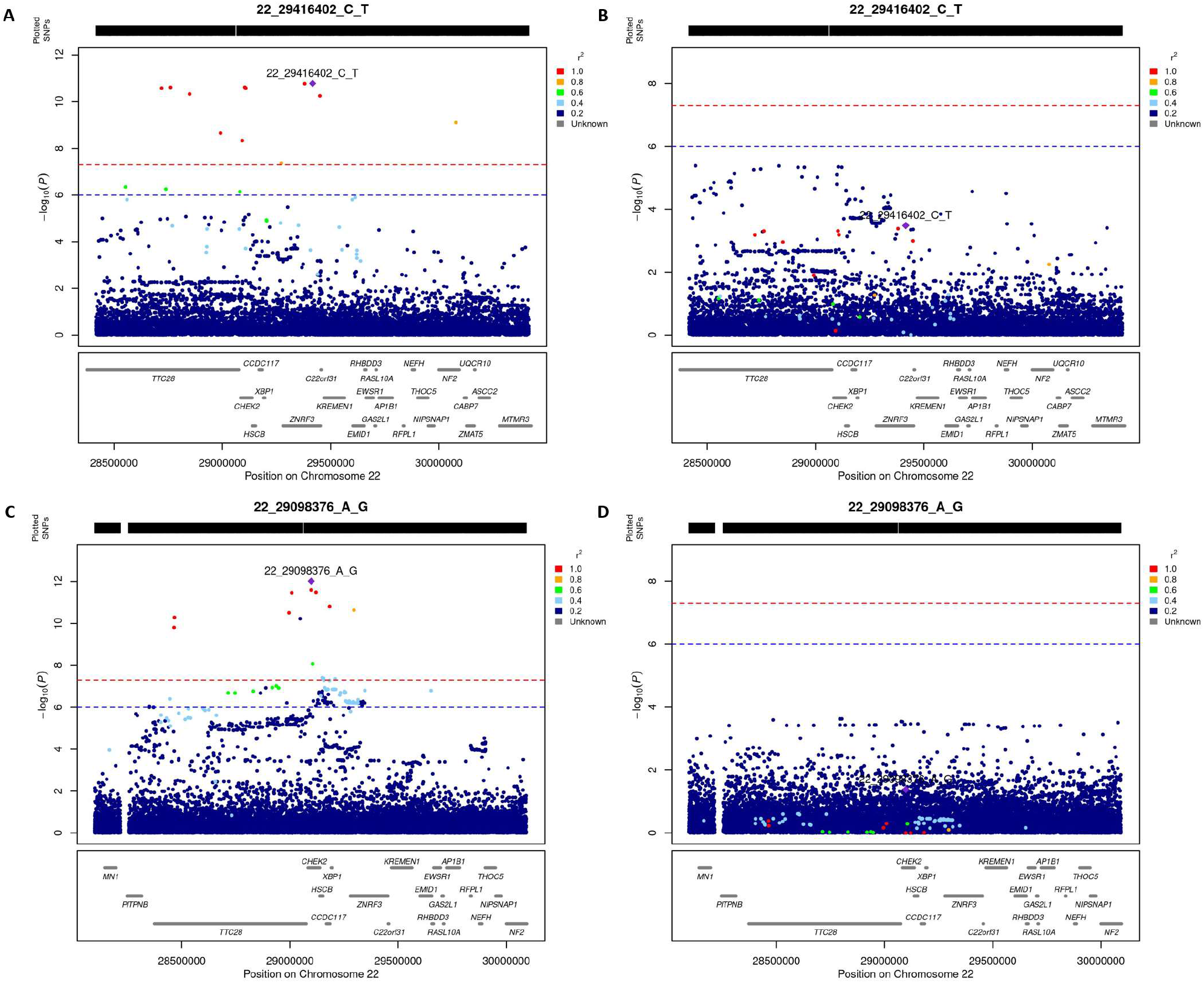
Regional plots before and after conditional analyses for lead variants in chromosome 22. FinnGen lead variant in locus 22q11 (A) along with conditional analysis results with frameshift variant (rs555607708) (B). Regional plot for the Estonian biobank lead variant in the same locus 22q11 before and after conditional analysis with linked missense variant (rs17879961) are shown in C and D. Regional plots were produced with R-package LocusZooms (https://github.com/Geeketics/LocusZooms/). r2 estimates were generated using LDstore [89] with SiSu v3 project WGS data consisting of 3775 individuals with Finnish ancestry.

When adjusting the GWAS for age and BMI, the FinnGen lead variant rs145598156 remained genome-wide significant [p = 4.5 × 10^−8^, OR= 13.5 (5.35-34.38)] (Table 1, Supplementary Fig 2).

When we tested for an interaction between PCOS, c.1100delC, and obesity using a logit regression model, a p-value of 0.066 for c.1100delC-BMI interaction was obtained (OR 1.04, 95 % CI 0.99-1.09).

### Validation GWAS detected an independent association in *CHEK2*

A validation GWAS performed in the EstBB also uncovered a genome-wide significant association [p=1.3 × 10^−12^, OR=1.64 (1.34-1.88)] in the 22q11 region. The lead variant rs182075939 was an intron variant located 22 kb from the TSS of *TTC28* (Fig 1B and 2C). Functional annotation revealed a tightly linked missense variant rs17879961 (r2=0.83, p=4.23 × 10^−12^), known as I157T, in *CHEK2*, which has been shown to alter CHEK2 ability to bind p53, BRCA1, and Cdc25A proteins [56,57]. EstBB lead variant rs182075939 presents a higher allele frequency in Estonians (AF=0.048) compared to Finns (AF=0.029) and other European populations (AF=0.017) (PMID: 32461654). The analysis conditioned on I157T resulted in no genome-wide significant associations in this locus, with a p-value of 0.04 for the lead variant rs182075939 (Fig 2D).

When adjusting the GWAS for age and BMI, EstBB lead variant rs182075939 remained genome-wide significant [p=4.6 × 10^−11^, OR=1.68 (1.44-1.96)] (Table 1, Supplementary Fig 2).

Interestingly, even though the association signals found in EstBB and FinnGen data sets overlap with each other (Fig 3), they seem to be part of independent haplotypes with r^2^-value below 0.05 between the lead variants. The lead variant of FinnGen data had a p-value of 0.031 in EstBB. The EstBB lead variant had a p-value of 1.8 × 10^−5^ in FinnGen (Table 1).

**Fig 3.**
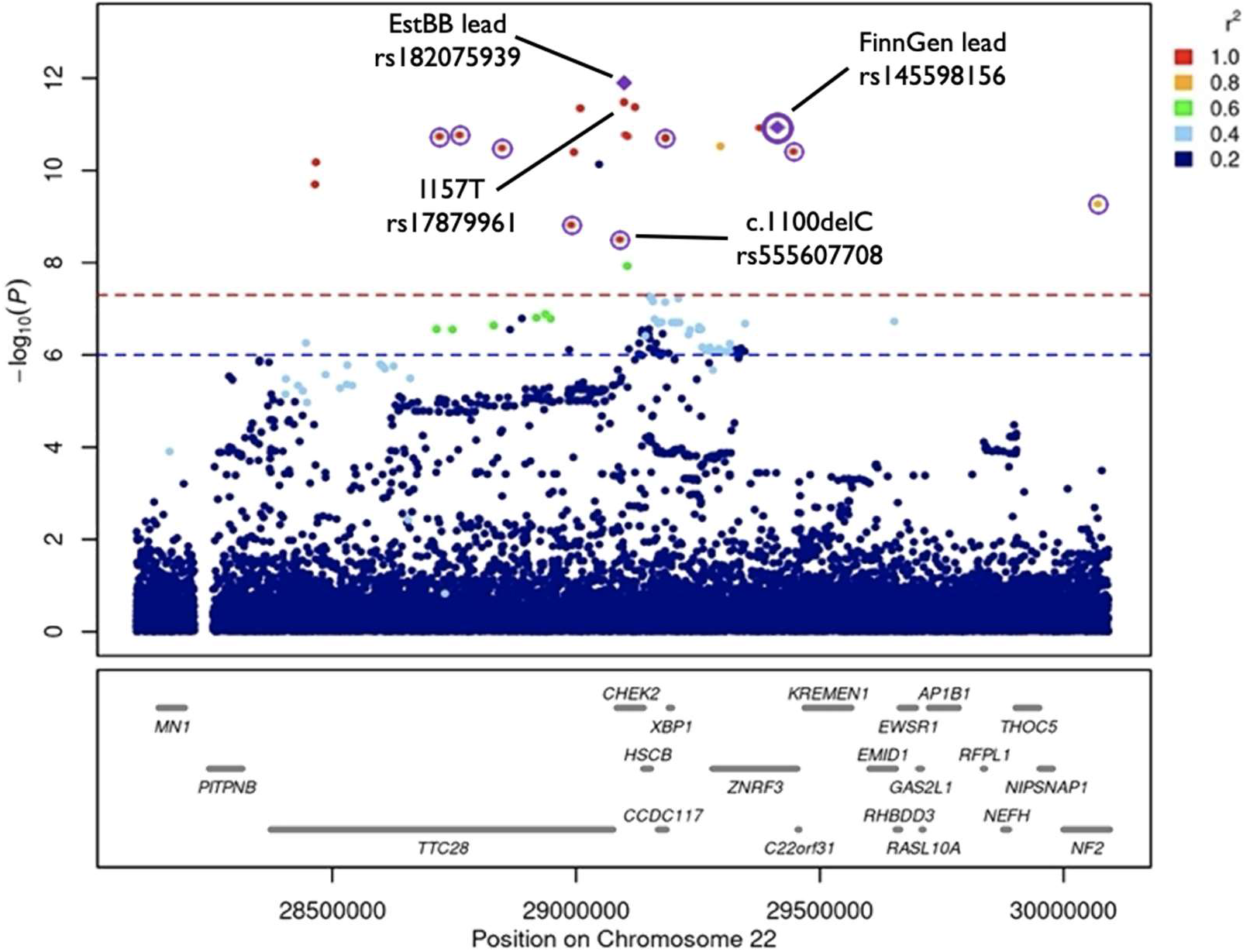
CHEK2 variants. Independent FinnGen and Estonian Biobank GWAS associations overlapping the CHEK2 gene are plotted on a single LocusZooms figure. Genome-wide significant variants in FinnGen data are denoted with purple circles; Estonian Biobank specific variants are not circled.

### Meta-analysis confirmed and expanded novel associations with PCOS in *CHEK2* and *MYO10*

A meta-analysis was performed for the two GWAS incorporating a total of 3,609 women with PCOS and 229,788 controls. In the meta-analysis, the FinnGen lead variant on chromosome 22 rs145598156 had a p-value of 3.6 × 10^−8^ with significant heterogeneity between cohorts (p^het^= 9.58 × 10^−6^), while EstBB lead variant rs182075939 showed a p-value of 1.9 × 10^−16^ in the meta-analyses results without significant heterogeneity between cohorts (p^het^=0.3). When the FinnGen and EstBB results were conditioned for the c.1100delC and I157T variants and the results were meta-analyzed, there were no additional genome-wide significant signals in the *CHEK2* locus.

The meta-analysis also revealed three more variants associating with PCOS, in addition to the three detected in FinnGen and EstBB GWAS separately (Table 1, Supplementary Fig 1). Two of the additional signals were in chromosome 11 and have been previously shown to be associated with PCOS: rs11031002 is located near *FSHB* and rs1672716 is an intron variant of *ZBTB16*. The third new association peak in the meta-analysis [rs9312937, p= 1.7 × 10^−8^, OR=1.16 (1.10-1.22), AF=0.44] was a common variant in an intronic region of chromosome 5, located 100 kb from the TSS of the *MYO10* gene, which to our knowledge has not previously been associated with PCOS. A total of two potentially causal genes were suggested by chromatin interaction data from 21 different tissues/cell types, with *MYO10* being the closest one, while no significant eQTL associations were detected using FUMA [46] in this locus.

The average effect sizes of the novel alleles described in chromosome 22 (OR= 1.69-3.01) (Table 1) were higher than the effects observed for alleles associated with PCOS in the rest of the common variants described (OR=1.06-1.40), which could be explained by the often-observed inverse relationship between allele frequency and effect size [58]. Moreover, we observed consistency in the direction of effects between the three datasets analyzed (discovery, validation, and joint meta-analysis) (Fig 4).

**Fig 4.**
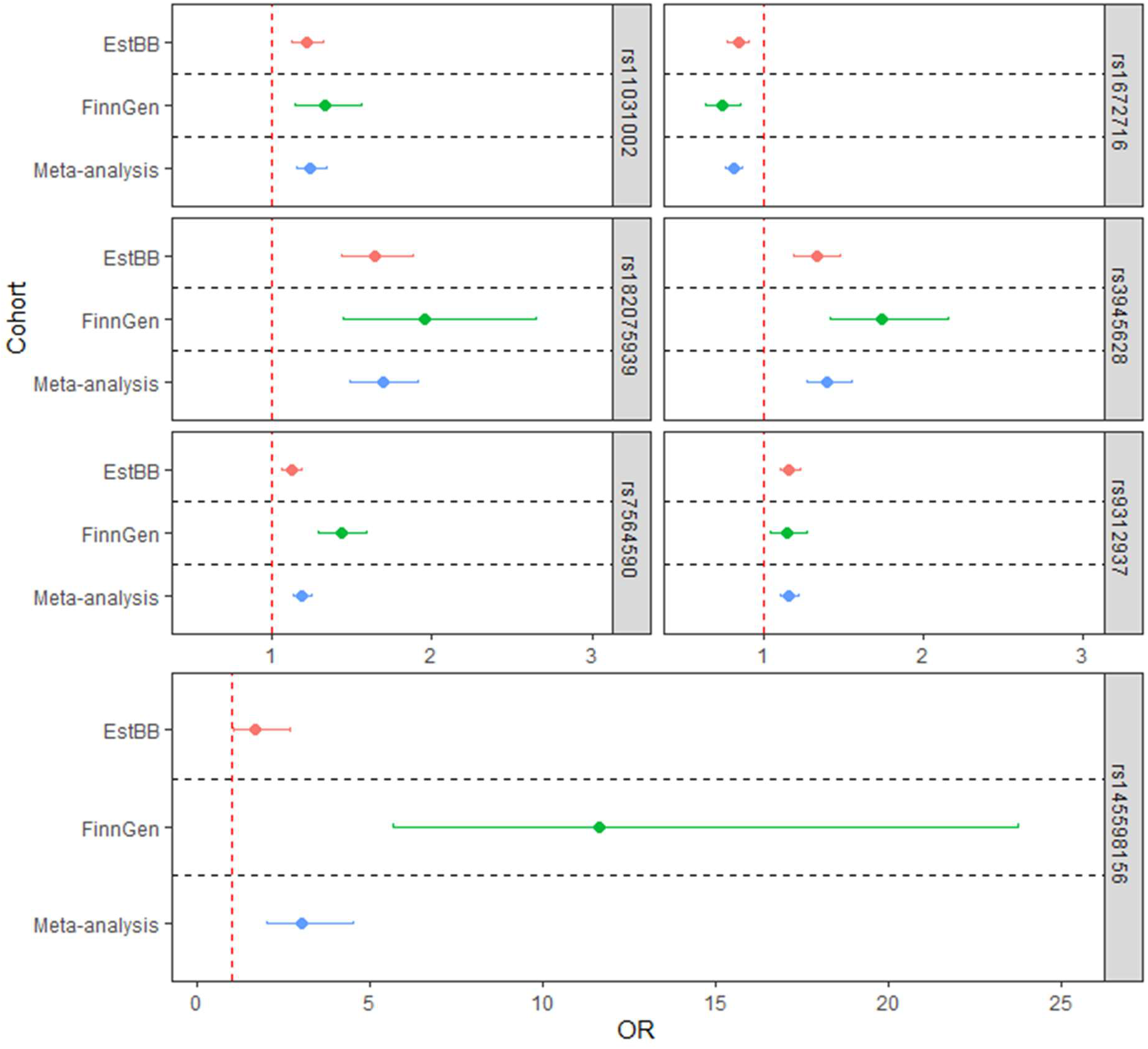
Forest plot of effect estimates for the seven lead variants associated with PCOS. The odds ratios (dots) and 95% confidence intervals (error bars) are shown for the two included cohorts and meta-analysis

In colocalization analyses, all posterior probabilities for a shared causal variant were lower than 0.8, thus we did not find enough evidence that two association signals in the genome-wide association analysis and gene expression are consistent with a shared causal variant.

### A testosterone-associated variant in intron of PLEKHM3 also associated with PCOS

We also utilized a set of 675 previously reported testosterone and SHBG related variants [54] to search for associations with PCOS, from which 60 were nominally significant in our meta-analysis results (Supplementary Table 1). Statistically, the most significant variant in our meta-analysis of this set was variant rs11031005 close to the FSHB (p= 2.74 × 10^−8^). However, we also found an additional association on chromosome 2 in an intronic region located 116 kb from the TSS of PLEKHM3 (rs873779, p= 1.7 × 10^−5^). According to the GTEx database (GTEx Consortium, PMID: 23715323), this variant is an eQTL for adjacent FZD5 expression in the esophagus (p=4.2 × 10^−8^), minor salivary gland (p=3.1 × 10^−6^), skin (4.5 × 10^−6^), and adrenal gland (p=2.7 × 10^−5^).

## Discussion

In this study, we found two independent novel associations for PCOS on 22q11.2. Both associations had tightly linked variants, a frameshift (c.1100delC), and a missense (I157T), in the *CHEK2* gene. A novel association was also detected in an intron of *MYO10*. We were also able to replicate signals commonly reported in PCOS GWAS – *DENND1A, ERBB4* (*HER4*), ZBTB16 and *FSHB* – in our North-European populations.

*CHEK2* rs555607708 (c.1100delC), the likely association driving variant in FinnGen, is a Finnish enriched variant with a 3.7-fold enrichment compared to non-Finnish, non-Estonian Europeans, with an enrichment of 1.7 compared to Estonians [59]. In a similar way, I157T, the likely association driving variant in EstBB, has a substantially higher allele frequency in the Estonian (0.048) and Finnish (0.029) populations, compared to the Non-Finnish, North-Western European population (0.002) according to the gnomAD database [55]. The enrichment of the alleles likely allowed us to detect the associations with PCOS in the Finnish and Estonian populations, whereas in populations with lower minor allele frequencies, much larger study populations would need to be used.

Checkpoint kinase 2 (CHEK2) is a mediator of DNA damage signaling in response to double-stranded (ds) DNA breaks. During the dsDNA damage, CHEK2 is activated, resulting in phosphorylation of proteins involved in DNA repair, cell cycle regulation, and apoptosis. Thus, CHEK2 can be considered an important factor in quality control of cells. If CHEK2 function is disturbed, DNA repair is imbalanced, which can lead to genomic instability and tumorigenesis [60]. Consequently, *CHEK2* variants have been associated with various cancers, particularly in breast cancer [61,62] but also in endometrial cancer [63]. Interestingly, the c.1100delC variant in *CHEK2* has been shown in an earlier study to particularly predispose obese carriers to development of breast cancer [53]. The interaction between BMI and PCOS associated variants has previously been suggested for example for the FTO alpha-ketoglutarate-dependent dioxygenase gene [64]. Although our results did not reach statistical significance to support this occurring in the case of c.1100delC, a replication of this analysis with a larger samples size is needed. Nevertheless, maintaining a healthy body weight seems to be advisable especially for carriers of c.1100delC.

Epidemiological studies have shown an increased risk for endometrial cancer in women with PCOS. However, this does not apply to other gynecological cancers like ovarian, cervical, or breast cancer [13-15,65,66]. It is important to recognize that association does not imply causation, and further studies are required to deduce a cause-and-effect relationship between these factors. In line with this, three very recently published studies utilizing the mendelian randomization approach have suggested a modest but significant causal effect between PCOS and breast cancer [67-69]. Nonetheless, the fact that the risks do not seem to translate into clinical findings is notable, and may indicate, for example, more efficient DNA repair systems in women with PCOS, which has also been associated with a later onset of menopause [27,70].

Interestingly, CHEK2 is also a critical DNA damage checkpoint protein in meiotic prophase I oocytes. CHEK2 activation plays a crucial role in fetal oocyte attrition, a phenomenon through which 80% of the initial ovarian oocyte reserve is lost during fetal development in mammals [71]. Deletion of *Chk2* in mice leads to a maximized ovarian reserve at postnatal day 2 (P2); however, prepubertal (P19) *Chk2-/-* mice have a comparable number of oocytes, and the number of litters and pups per litter are comparable to *Chk2+/*-littermates in adult mice [71]. Nevertheless, at 13.5 months *Chk2-/-* mice have reduced follicle atresia, a higher number of ovulated MII oocytes, and higher AMH levels [70]. In the same study, it was also reported that a *CHEK2* loss-of-function allele is associated with later menopausal age in humans [70]. This would be in line with women with PCOS, as they also present with an increased ovarian reserve, higher AMH levels even at later reproductive years, and delayed menopause [5-7,9,72]. A specific association between menopause delaying alleles and PCOS has also been previously demonstrated [26]. In a recent preprint work Ward et al found that *CHEK2* was associated with age of menopause. When conducting a phenome wide association study (PheWAS) on their associations, an aggregate of all *CHEK2* damaging variants also showed an association with PCOS, in line with our findings [74].

Our study also adds support to previously reported associations with PCOS near or in genes such as *ERBB4, DENND1A, FSHB*, and *ZBTB16*. Interestingly, *ERBB4* has also recently been linked to follicle development, as Veikkolainen *et al*. showed that after a specific conditional knock-out of *Erbb4* in granulosa cells, the mice presented a PCOS-like phenotype with arrested follicle development, subfertility, hyperandrogenism, high luteinizing hormone secretion, and high AMH levels in the ovarian follicles and circulation. The mice were obese and showed metabolic dysfunction as well as increased insulin secretion. *ERBB4* appears to be essential for proper oocyte maturation and ovulation [75]. Thus, the present study reinforces the link between PCOS and abnormal follicle development and high levels of AMH.

Although the role of novel common variants will probably be better captured in studies of larger sample sizes regardless of larger genetic variation, this study presents an interesting novel association in an intronic region of *MYO10*. The *MYO10* gene codes for an atypical myosin, which is involved in filopodia formation, phagocytosis, and cargo transport in cells [76]. Genetic variation in *MYO10* has previously been linked to type 2 diabetes [77] and traits of metabolic syndrome [78]. Interestingly, the variant we identified here is also associated with the age at menarche [79], which supports a plausible role in reproduction. Although a metabolic link between *MYO10* and PCOS seems likely, further research is needed to characterize the role of *MYO10* in PCOS.

When we compared our significant variants to a set of candidate variants associated with high levels of total testosterone, bioavailable testosterone, or SHGB levels in women in a recent study, a new association with PCOS was found in the intronic region of *PLEKHM3* (rs873779, p=1.7 × 10^−5^). However, this variant has been reported to act as an eQTL that modifies the expression *FZD5*, a Wnt receptor protein, in multiple tissues [80]. Proper Wnt signaling is important for ovarian development and oocyte maturation [81,82] and abnormalities in this pathway have been reported in the endometrium, granulosa cells, and adipose tissue of women with PCOS [83-85].

As previous studies have suggested that obesity may have a causal role in PCOS [39,40] we reran the association analyses adjusting for BMI. A reduction in significance of several associations was expected due to limited availability of BMI measurement data in the sampled individuals (60% in Finngen and 75% in EstBB). The two replicated (*FSHB, ZBTB16)* and the two novel associations near *MYO10* and *CHEK2* fell below genome-wide significance. It is therefore difficult to deduce whether this was due to BMI adjustment or loss of power. Thus, we mainly focused on age-adjusted associations and we acknowledge that larger sample sizes are needed to further explore the interplay between BMI and PCOS genetic factors.

Overall, it is important to note that complex LD patterns between association signals might eclipse more distant causal genes. In order to infer plausible shared causal variants between PCOS genetic variants and gene expression, we conducted colocalization analyses, which did not show any significant findings. This might be explained by the reduced sample size in gene expression panels that study tissues of interest in PCOS such as reproductive tissues, which might result in a lack of power to detect any significant associations. Thus, further functional studies are needed to better characterize the regulatory functions of the loci uncovered.

Our main discovery of the two rare variants near *CHEK2* that influence PCOS underlines the value of using study populations with a distinct genetic makeup. The demographic history can have a profound effect on the allele frequency spectrum of a population and may result in locally varying genetic architectures for medical conditions. Interplay of past events such as population bottlenecks, founder effects, range expansions, and genetic drift may increase the relative frequency of many clinically important genetic variants [33,86]. In such populations, selection may not have had sufficient time to reduce their frequencies and the strength of the selection can be reduced, especially in populations with small effective sizes. When such enriched (or even private) alleles are causal or linked to causal variation, an increased statistical power is present, enabling their detection in association analysis, thus the discovery of new candidate genes would be favored, as shown in earlier studies [87,88]. Furthermore, isolated populations tend to have uniform environment and it is also often easier to standardize phenotype definitions [86].

The main strength of this study was the use of the two large, comprehensive genetic data sets, FinnGen and EstBB, which have been extensively linked to national registers, such as The Care Register for Health Care in Finland and the Estonian Health Insurance Fund registries in Estonia, and with other relevant databases [42]. Both populations are genetically well characterized and the Finnish population is isolated due to serial founder effects and limited gene flow in the past [89].

The register-based approach is also a limiting factor, as the health register-based prevalence of PCOS is very low our study populations, plausibly reflecting underdiagnosis of the syndrome. Nevertheless, we were able to replicate four previously reported signals, *ERBB4, DENND1A, FSHB* and *ZBTB16*, which adds reliability to our results. Given the register-based approach, we were not able to assess in more detail the different PCOS phenotypes; however, a previous study indicated that women with PCOS diagnosed by a physician using different diagnostic criteria are genetically similar [27]. Further experimental work is also warranted to confirm and strengthen the role of the candidate genes we propose in the identified loci, but the current findings will pave way for these studies.

In conclusion, we identified two rare population-enriched variants located in *CHEK2* that are significantly associated with PCOS. We also supported and expanded on previous knowledge of the association between common genetic variants and the disorder. These findings emphasize the importance of including and studying unique populations such as those of Finland and Estonia when performing genetic studies of complex diseases and to advance our understanding of genetic factors underlying PCOS.

## Supporting information

Supplementary Checklist 1

## Data Availability

Full meta-analysis summary statistics will be made available upon publication.

## Acknowledgements

We thank all FinnGen and EstBB participants for offering us the valuable resources. We also acknowledge the Estonian Biobank Research team members Andres Metspalu, Tõnu Esko, Mari Nelis and Lili Milani. This work has received funding from the European Union’s Horizon 2020 research and innovation programme under the Marie Skłodowska-Curie grant agreement No 813707 (N.P.G, T.L., T.P.), the Estonian Research Council grant (PRG687, T.L.), the Academy of Finland grants 315921 (T.P.), 321763 (T.P.), 297338 (J.K.), 307247 (J.K.), 344695 (H.L.), Novo Nordisk Foundation grant NNF17OC0026062 (J.K.), the Sigrid Juselius Foundation project grants (T.L, J.K.), Finska Läkaresällskapet (H.L.) and Jane and Aatos Erkko Foundation (H.L). The funders had no role in study design, data collection and analysis, decision to publish, or preparation of the manuscript.

## Supplementary information

Supplementary Note 1: Contributors of FinnGen.

Supplementary Checklist 1: STREGA (STrengthening the REporting of Genetic Association studies (STREGA) reporting recommendations) report.

Supplementary Table 1: Look-up of the markers previously associated with A) testosterone, B) SHBG and C) bioavailable testosterone.

Supplementary Fig 1: Regional association plots of genome-wide significant variants.

Supplementary Fig 2: Manhattan plot for age-and BMI-adjusted GWAS in the Finnish dataset, the Estonian dataset GWAS and the joint GWAS meta-analysis of PCOS.

