## Supplementary Note 1 for "Leveraging Northern European population history; novel low frequency variants for polycystic ovary syndrome"

### Contributors of FinnGen


#### Steering Committee

Aarno Palotie Institute for Molecular Medicine Finland, HiLIFE, University of Helsinki, Finland

Mark Daly Institute for Molecular Medicine Finland, HiLIFE, University of Helsinki, Finland

**Pharmaceutical companies**

Bridget Riley-Gills Abbvie, Chicago, IL, United States

Howard Jacob Abbvie, Chicago, IL, United States

Dirk Paul Astra Zeneca, Cambridge, United Kingdom

Heiko Runz Biogen, Cambridge, MA, United States

Sally John Biogen, Cambridge, MA, United States

Robert Plenge Celgene, Summit, NJ, United States/Bristol Myers Squibb, New York, NY, United States

Mark McCarthy Genentech, San Francisco, CA, United States

Julie Hunkapiller Genentech, San Francisco, CA, United States

Meg Ehm GlaxoSmithKline, Brentford, United Kingdom

Kirsi Auro GlaxoSmithKline, Brentford, United Kingdom

Caroline Fox Merck, Kenilworth, NJ, United States

Anders Mälarstig Pfizer, New York, NY, United States

Katherine Klinger Sanofi, Paris, France

Deepak Raipal Sanofi, Paris, France

Tim Behrens Maze Therapeutics, San Francisco, CA, United States

Robert Yang Janssen Biotech, Beerse, Belgium

Richard Siegel Novartis, Basel, Switzerland

**University of Helsinki & Biobanks**

Tomi Mäkelä HiLIFE, University of Helsinki, Finland, Finland

Jaakko Kaprio Institute for Molecular Medicine Finland, HiLIFE, Helsinki, Finland, Finland

Petri Virolainen Auria Biobank / University of Turku / Hospital District of Southwest Finland, Turku, Finland

Antti Hakanen Auria Biobank / University of Turku / Hospital District of Southwest Finland, Turku, Finland

Terhi Kilpi THL Biobank / The National Institute of Health and Welfare Helsinki, Finland

Markus Perola THL Biobank / The National Institute of Health and Welfare Helsinki, Finland

Jukka Partanen Finnish Red Cross Blood Service / Finnish Hematology Registry and Clinical Biobank, Helsinki, Finland

Anne Pitkäranta Helsinki Biobank / Helsinki University and Hospital District of Helsinki and Uusimaa, Helsinki

Juhani Junttila Northern Finland Biobank Borealis / University of Oulu / Northern Ostrobothnia Hospital District, Oulu, Finland

Raisa Serpi Northern Finland Biobank Borealis / University of Oulu / Northern Ostrobothnia Hospital District, Oulu, Finland

Tarja Laitinen Finnish Clinical Biobank Tampere / University of Tampere / Pirkanmaa Hospital District, Tampere, Finland

Johanna Mäkelä Finnish Clinical Biobank Tampere / University of Tampere / Pirkanmaa Hospital District, Tampere, Finland

Veli-Matti Kosma Biobank of Eastern Finland / University of Eastern Finland / Northern Savo Hospital District, Kuopio, Finland

Urho Kujala Central Finland Biobank / University of Jyväskylä / Central Finland Health Care District, Jyväskylä, Finland

**Other Experts/ Non-Voting Members**

Outi Tuovila Business Finland, Helsinki, Finland

Raimo Pakkanen Business Finland, Helsinki, Finland

#### Scientific Committee

**Pharmaceutical companies**

Jeffrey Waring Abbvie, Chicago, IL, United States

Ali Abbasi Abbvie, Chicago, IL, United States

Mengzhen Liu Abbvie, Chicago, IL, United States

Ioanna Tachmazidou Astra Zeneca, Cambridge, United Kingdom

Chia-Yen Chen Biogen, Cambridge, MA, United States

Heiko Runz Biogen, Cambridge, MA, United States

Shameek Biswas Celgene, Summit, NJ, United States/Bristol Myers Squibb, New York, NY, United States

Julie Hunkapiller Genentech, San Francisco, CA, United States

Meg Ehm GlaxoSmithKline, Brentford, United Kingdom

Neha Raghavan Merck, Kenilworth, NJ, United States

Adriana Huertas-Vazquez Merck, Kenilworth, NJ, United States

Anders Mälarstig Pfizer, New York, NY, United States

Xinli Hu Pfizer, New York, NY, United States

Katherine Klinger Sanofi, Paris, France

Matthias Gossel Sanofi, Paris, France

Robert Graham Maze Therapeutics, San Francisco, CA, United States

Tim Behrens Maze Therapeutics, San Francisco, CA, United States

Beryl Cummings Maze Therapeutics, San Francisco, CA, United States

Wilco Fleuren Janssen Biotech, Beerse, Belgium

Dawn Waterworth Janssen Biotech, Beerse, Belgium

Nicole Renaud Novartis, Basel, Switzerland

Aviv Madar Novartis, Basel, Switzerland

Maen Obeidat Novartis, Basel, Switzerland

**University of Helsinki & Biobanks**

Samuli Ripatti Institute for Molecular Medicine Finland, HiLIFE, Helsinki, Finland

Johanna Schleutker Auria Biobank / Univ. of Turku / Hospital District of Southwest Finland, Turku, Finland

Markus Perola THL Biobank / The National Institute of Health and Welfare Helsinki, Finland

Mikko Arvas Finnish Red Cross Blood Service / Finnish Hematology Registry and Clinical Biobank, Helsinki, Finland

Olli Carpén Helsinki Biobank / Helsinki University and Hospital District of Helsinki and Uusimaa, Helsinki

Reetta Hinttala Northern Finland Biobank Borealis / University of Oulu / Northern Ostrobothnia Hospital District, Oulu, Finland

Johannes Kettunen Northern Finland Biobank Borealis / University of Oulu / Northern Ostrobothnia Hospital District, Oulu, Finland

Johanna Mäkelä Finnish Clinical Biobank Tampere / University of Tampere / Pirkanmaa Hospital District, Tampere, Finland

Arto Mannermaa Biobank of Eastern Finland / University of Eastern Finland / Northern Savo Hospital District, Kuopio, Finland

Jari Laukkanen Central Finland Biobank / University of Jyväskylä / Central Finland Health Care District, Jyväskylä, Finland

Urho Kujala Central Finland Biobank / University of Jyväskylä / Central Finland Health Care District, Jyväskylä, Finland

#### Clinical Groups

**Neurology Group**

| | |
|---|---|
| Reetta Kälviäinen | Northern Savo Hospital District, Kuopio, Finland |
| Valtteri Julkunen | Northern Savo Hospital District, Kuopio, Finland |
| Hilkka Soininen | Northern Savo Hospital District, Kuopio, Finland |
| Anne Remes | Northern Ostrobothnia Hospital District, Oulu, Finland |
| Mikko Hiltunen | Northern Savo Hospital District, Kuopio, Finland |
| Jukka Peltola | Pirkanmaa Hospital District, Tampere, Finland |
| Pentti Tienari | Hospital District of Helsinki and Uusimaa, Helsinki, Finland |
| Juha Rinne | Hospital District of Southwest Finland, Turku, Finland |
| Roosa Kallionpää | Hospital District of Southwest Finland, Turku, Finland |
| Ali Abbasi | Abbvie, Chicago, IL, United States |
| Adam Ziemann | Abbvie, Chicago, IL, United States |
| Jeffrey Waring | Abbvie, Chicago, IL, United States |
| Sahar Esmaeeli | Abbvie, Chicago, IL, United States |
| Nizar Smaoui | Abbvie, Chicago, IL, United States |
| Anne Lehtonen | Abbvie, Chicago, IL, United States |
| Susan Eaton | Biogen, Cambridge, MA, United States |
| Heiko Runz | Biogen, Cambridge, MA, United States |

| | |
|---|---|
| Sanni Lahdenperä | Biogen, Cambridge, MA, United States |
| Janet van Adelsberg | Celgene, Summit, NJ, United States/ Bristol Myers Squibb, New York, NY, United States |
| Shameek Biswas | Celgene, Summit, NJ, United States/ Bristol Myers Squibb, New York, NY, United States |
| Julie Hunkapiller | Genentech, San Francisco, CA, United States |
| Natalie Bowers | Genentech, San Francisco, CA, United States |
| Edmond Teng | Genentech, San Francisco, CA, United States |
| Sarah Pendergrass | Genentech, San Francisco, CA, United States |
| Onuralp Soylemez | Merck, Kenilworth, NJ, United States |
| Kari Linden | Pfizer, New York, NY, United States |
| Fanli Xu | GlaxoSmithKline, Brentford, United Kingdom |
| David Pulford | GlaxoSmithKline, Brentford, United Kingdom |
| Kirsi Auro | GlaxoSmithKline, Brentford, United Kingdom |
| Laura Addis | GlaxoSmithKline, Brentford, United Kingdom |
| John Eicher | GlaxoSmithKline, Brentford, United Kingdom |
| Minna Raivio | Hospital District of Helsinki and Uusimaa, Helsinki, Finland |
| Sarah Pendergrass | Genentech, San Francisco, CA, United States |
| Beryl Cummings | Maze Therapeutics, San Francisco, CA, United States |
| Juulia Partanen | Institute for Molecular Medicine Finland, HiLIFE, University of Helsinki, Finland |

**Gastroenterology Group**

| | |
|---|---|
| Martti Färkkilä | Hospital District of Helsinki and Uusimaa, Helsinki, Finland |
| Jukka Koskela | Hospital District of Helsinki and Uusimaa, Helsinki, Finland |
| Sampsa Pikkarainen | Hospital District of Helsinki and Uusimaa, Helsinki, Finland |
| Airi Jussila | Pirkanmaa Hospital District, Tampere, Finland |
| Katri Kaukinen | Pirkanmaa Hospital District, Tampere, Finland |
| Timo Blomster | Northern Ostrobothnia Hospital District, Oulu, Finland |
| Mikko Kiviniemi | Northern Savo Hospital District, Kuopio, Finland |
| Markku Voutilainen | Hospital District of Southwest Finland, Turku, Finland |
| Ali Abbasi | Abbvie, Chicago, IL, United States |
| Graham Heap | Abbvie, Chicago, IL, United States |

| | |
|---|---|
| Jeffrey Waring | Abbvie, Chicago, IL, United States |
| Nizar Smaoui | Abbvie, Chicago, IL, United States |
| Fedik Rahimov | Abbvie, Chicago, IL, United States |
| Anne Lehtonen | Abbvie, Chicago, IL, United States |
| Keith Usiskin | Celgene, Summit, NJ, United States/ Bristol Myers Squibb, New York, NY, United States |
| Tim Lu | Genentech, San Francisco, CA, United States |
| Natalie Bowers | Genentech, San Francisco, CA, United States |
| Danny Oh | Genentech, San Francisco, CA, United States |
| Sarah Pendergrass | Genentech, San Francisco, CA, United States |
| Kirsi Kalpala | Pfizer, New York, NY, United States |
| Melissa Miller | Pfizer, New York, NY, United States |
| Xinli Hu | Pfizer, New York, NY, United States |
| Linda McCarthy | GlaxoSmithKline, Brentford, United Kingdom |
| Onuralp Soylemez | Merck, Kenilworth, NJ, United States |
| Mark Daly | Institute for Molecular Medicine Finland, HiLIFE, University of Helsinki, Finland |


**Rheumatology Group**


| | |
|---|---|
| Kari Eklund | Hospital District of Helsinki and Uusimaa, Helsinki, Finland |
| Antti Palomäki | Hospital District of Southwest Finland, Turku, Finland |
| Pia Isomäki | Pirkanmaa Hospital District, Tampere, Finland |
| Laura Pirilä | Hospital District of Southwest Finland, Turku, Finland |
| Oili Kaipiainen-Seppänen | Northern Savo Hospital District, Kuopio, Finland |
| Johanna Huhtakangas | Northern Ostrobothnia Hospital District, Oulu, Finland |
| Ali Abbasi | Abbvie, Chicago, IL, United States |
| Jeffrey Waring | Abbvie, Chicago, IL, United States |
| Fedik Rahimov | Abbvie, Chicago, IL, United States |
| Apinya Lertratanakul | Abbvie, Chicago, IL, United States |
| Nizar Smaoui | Abbvie, Chicago, IL, United States |
| Anne Lehtonen | Abbvie, Chicago, IL, United States |
| David Close | Astra Zeneca, Cambridge, United Kingdom |

| | |
|---|---|
| Marla Hochfeld | Celgene, Summit, NJ, United States/ Bristol Myers Squibb, New York, NY, United States |
| Natalie Bowers | Genentech, San Francisco, CA, United States |
| Sarah Pendergrass | Genentech, San Francisco, CA, United States |
| Onuralp Soylemez | Merck, Kenilworth, NJ, United States |
| Kirsi Kalpala | Pfizer, New York, NY, United States |
| Nan Bing | Pfizer, New York, NY, United States |
| Xinli Hu | Pfizer, New York, NY, United States |
| Jorge Esparza Gordillo | GlaxoSmithKline, Brentford, United Kingdom |
| Kirsi Auro | GlaxoSmithKline, Brentford, United Kingdom |
| Dawn Waterworth | Janssen Biotech, Beerse, Belgium |
| Nina Mars | Institute for Molecular Medicine Finland, HiLIFE, Helsinki, Finland |

**Pulmonology Group**

| | |
|---|---|
| Tarja Laitinen | Pirkanmaa Hospital District, Tampere, Finland |
| Margit Pelkonen | Northern Savo Hospital District, Kuopio, Finland |
| Paula Kauppi | Hospital District of Helsinki and Uusimaa, Helsinki, Finland |
| Hannu Kankaanranta | Pirkanmaa Hospital District, Tampere, Finland |
| Terttu Harju | Northern Ostrobothnia Hospital District, Oulu, Finland |
| Riitta Lahesmaa | Hospital District of Southwest Finland, Turku, Finland |
| Nizar Smaoui | Abbvie, Chicago, IL, United States |
| Alex Mackay | Astra Zeneca, Cambridge, United Kingdom |
| Glenda Lassi | Astra Zeneca, Cambridge, United Kingdom |
| Susan Eaton | Biogen, Cambridge, MA, United States |
| Steven Greenberg | Celgene, Summit, NJ, United States/ Bristol Myers Squibb, New York, NY, United States |
| Hubert Chen | Genentech, San Francisco, CA, United States |
| Sarah Pendergrass | Genentech, San Francisco, CA, United States |
| Natalie Bowers | Genentech, San Francisco, CA, United States |
| Joanna Betts | GlaxoSmithKline, Brentford, United Kingdom |
| Soumitra Ghosh | GlaxoSmithKline, Brentford, United Kingdom |
| Kirsi Auro | GlaxoSmithKline, Brentford, United Kingdom |

| | |
|---|---|
| Rajashree Mishra | GlaxoSmithKline, Brentford, United Kingdom |
| Sina Rüeger | Institute for Molecular Medicine Finland, HiLIFE, University of Helsinki, Finland |


**Cardiometabolic Diseases Group**

| | |
|---|---|
| Teemu Niiranen | The National Institute of Health and Welfare Helsinki, Finland |
| Felix Vaura | The National Institute of Health and Welfare Helsinki, Finland |
| Veikko Salomaa | The National Institute of Health and Welfare Helsinki, Finland |
| Markus Juonala | Hospital District of Southwest Finland, Turku, Finland |
| Kaj Metsärinne | Hospital District of Southwest Finland, Turku, Finland |
| Mika Kähönen | Pirkanmaa Hospital District, Tampere, Finland |
| Juhani Junttila | Northern Ostrobothnia Hospital District, Oulu, Finland |
| Markku Laakso | Northern Savo Hospital District, Kuopio, Finland |
| Jussi Pihlajamäki | Northern Savo Hospital District, Kuopio, Finland |
| Daniel Gordin | Hospital District of Helsinki and Uusimaa, Helsinki, Finland |
| Juha Sinisalo | Hospital District of Helsinki and Uusimaa, Helsinki, Finland |
| Marja-Riitta Taskinen | Hospital District of Helsinki and Uusimaa, Helsinki, Finland |
| Tiinamaija Tuomi | Hospital District of Helsinki and Uusimaa, Helsinki, Finland |
| Jari Laukkanen | Central Finland Health Care District, Jyväskylä, Finland |
| Benjamin Challis | Astra Zeneca, Cambridge, United Kingdom |
| Dirk Paul | Astra Zeneca, Cambridge, United Kingdom |
| Julie Hunkapiller | Genentech, San Francisco, CA, United States |
| Natalie Bowers | Genentech, San Francisco, CA, United States |
| Sarah Pendergrass | Genentech, San Francisco, CA, United States |
| Onuralp Soylemez | Merck, Kenilworth, NJ, United States |
| Jaakko Parkkinen | Pfizer, New York, NY, United States |
| Melissa Miller | Pfizer, New York, NY, United States |
| Russell Miller | Pfizer, New York, NY, United States |
| Audrey Chu | GlaxoSmithKline, Brentford, United Kingdom |
| Kirsi Auro | GlaxoSmithKline, Brentford, United Kingdom |
| Keith Usiskin | Celgene, Summit, NJ, United States/ Bristol Myers Squibb, New York, NY, United States |

Amanda Elliott Institute for Molecular Medicine Finland, HiLIFE, University of Helsinki, Finland / Broad Institute, Cambridge, MA, United States

Joel Rämö Institute for Molecular Medicine Finland, HiLIFE, University of Helsinki, Finland

Samuli Ripatti Institute for Molecular Medicine Finland, HiLIFE, University of Helsinki, Finland

Mary Pat Reeve Institute for Molecular Medicine Finland, HiLIFE, University of Helsinki, Finland

Sanni Ruotsalainen Institute for Molecular Medicine Finland, HiLIFE, University of Helsinki, Finland

**Oncology Group**

Tuomo Meretoja Hospital District of Helsinki and Uusimaa, Helsinki, Finland

Heikki Joensuu Hospital District of Helsinki and Uusimaa, Helsinki, Finland

Olli Carpén Hospital District of Helsinki and Uusimaa, Helsinki, Finland

Lauri Aaltonen Hospital District of Helsinki and Uusimaa, Helsinki, Finland

Johanna Mattson Hospital District of Helsinki and Uusimaa, Helsinki, Finland

Annika Auranen Pirkanmaa Hospital District , Tampere, Finland

Peeter Karihtala Northern Ostrobothnia Hospital District, Oulu, Finland

Saila Kauppila Northern Ostrobothnia Hospital District, Oulu, Finland

Päivi Auvinen Northern Savo Hospital District, Kuopio, Finland

Klaus Elenius Hospital District of Southwest Finland, Turku, Finland

Johanna Schleutker Hospital District of Southwest Finland, Turku, Finland

Relja Popovic Abbvie, Chicago, IL, United States

Jeffrey Waring Abbvie, Chicago, IL, United States

Bridget Riley-Gillis Abbvie, Chicago, IL, United States

Anne Lehtonen Abbvie, Chicago, IL, United States

Jennifer Schutzman Genentech, San Francisco, CA, United States

Julie Hunkapiller Genentech, San Francisco, CA, United States

Natalie Bowers Genentech, San Francisco, CA, United States

Sarah Pendergrass Genentech, San Francisco, CA, United States

Andrey Loboda Merck, Kenilworth, NJ, United States

Aparna Chhibber Merck, Kenilworth, NJ, United States

Heli Lehtonen Pfizer, New York, NY, United States

| | |
|---|---|
| Stefan McDonough | Pfizer, New York, NY, United States |
| Marika Crohns | Sanofi, Paris, France |
| Sauli Vuoti | Sanofi, Paris, France |
| Diptee Kulkarni | GlaxoSmithKline, Brentford, United Kingdom |
| Kirsi Auro | GlaxoSmithKline, Brentford, United Kingdom |
| Esa Pitkänen | Institute for Molecular Medicine Finland, HiLIFE, University of Helsinki, Finland |
| Nina Mars | Institute for Molecular Medicine Finland, HiLIFE, University of Helsinki, Finland |
| Mark Daly | Institute for Molecular Medicine Finland, HiLIFE, University of Helsinki, Finland |

**Opthalmology Group**

| | |
|---|---|
| Kai Kaarniranta | Northern Savo Hospital District, Kuopio, Finland |
| Joni A Turunen | Hospital District of Helsinki and Uusimaa, Helsinki, Finland |
| Terhi Ollila | Hospital District of Helsinki and Uusimaa, Helsinki, Finland |
| Sanna Seitsonen | Hospital District of Helsinki and Uusimaa, Helsinki, Finland |
| Hannu Uusitalo | Pirkanmaa Hospital District, Tampere, Finland |
| Vesa Aaltonen | Hospital District of Southwest Finland, Turku, Finland |
| Hannele Uusitalo-Järvinen | Pirkanmaa Hospital District, Tampere, Finland |
| Marja Luodonpää | Northern Ostrobothnia Hospital District, Oulu, Finland |
| Nina Hautala | Northern Ostrobothnia Hospital District, Oulu, Finland |
| Mengzhen Liu | Abbvie, Chicago, IL, United States |
| Heiko Runz | Biogen, Cambridge, MA, United States |
| Stephanie Loomis | Biogen, Cambridge, MA, United States |
| Erich Strauss | Genentech, San Francisco, CA, United States |
| Natalie Bowers | Genentech, San Francisco, CA, United States |
| Hao Chen | Genentech, San Francisco, CA, United States |
| Sarah Pendergrass | Genentech, San Francisco, CA, United States |
| Anna Podgornaia | Merck, Kenilworth, NJ, United States |
| Juha Karjalainen | Institute for Molecular Medicine Finland, HiLIFE, University of Helsinki, Finland / Broad Institute, Cambridge, MA, United States |
| Esa Pitkänen | Institute for Molecular Medicine Finland, HiLIFE, University of Helsinki, Finland |

**Dermatology Group**

| | |
|---|---|
| Kaisa Tasanen | Northern Ostrobothnia Hospital District, Oulu, Finland |
| Laura Huilaja | Northern Ostrobothnia Hospital District, Oulu, Finland |
| Katariina Hannula-Jouppi | Hospital District of Helsinki and Uusimaa, Helsinki, Finland |
| Teea Salmi | Pirkanmaa Hospital District, Tampere, Finland |
| Sirkku Peltonen | Hospital District of Southwest Finland, Turku, Finland |
| Leena Koulu | Hospital District of Southwest Finland, Turku, Finland |
| Kirsi Kalpala | Pfizer, New York, NY, United States |
| Ying Wu | Pfizer, New York, NY, United States |
| David Choy | Genentech, San Francisco, CA, United States |
| Sarah Pendergrass | Genentech, San Francisco, CA, United States |
| Nizar Smaoui | Abbvie, Chicago, IL, United States |
| Fedik Rahimov | Abbvie, Chicago, IL, United States |
| Anne Lehtonen | Abbvie, Chicago, IL, United States |
| Dawn Waterworth | Janssen Biotech, Beerse, Belgium |

**Odontology Group**

| | |
|---|---|
| Pirkko Pussinen | Hospital District of Helsinki and Uusimaa, Helsinki, Finland |
| Aino Salminen | Hospital District of Helsinki and Uusimaa, Helsinki, Finland |
| Tuula Salo | Hospital District of Helsinki and Uusimaa, Helsinki, Finland |
| David Rice | Hospital District of Helsinki and Uusimaa, Helsinki, Finland |
| Pekka Nieminen | Hospital District of Helsinki and Uusimaa, Helsinki, Finland |
| Ulla Palotie | Hospital District of Helsinki and Uusimaa, Helsinki, Finland |
| Juha Sinisalo | Hospital District of Helsinki and Uusimaa, Helsinki, Finland |
| Maria Siponen | Northern Savo Hospital District, Kuopio, Finland |
| Liisa Suominen | Northern Savo Hospital District, Kuopio, Finland |
| Päivi Mäntylä | Northern Savo Hospital District, Kuopio, Finland |
| Ulvi Gursoy | Hospital District of Southwest Finland, Turku, Finland |
| Vuokko Anttonen | Northern Ostrobothnia Hospital District, Oulu, Finland |

| | |
|---|---|
| Kirsi Sipilä | Northern Ostrobothnia Hospital District, Oulu, Finland |
| Sarah Pendergrass | Genentech, San Francisco, CA, United States |

**Women's Health and Reproduction Group**

| | |
|---|---|
| Hannele Laivuori | Institute for Molecular Medicine Finland, HiLIFE, University of Helsinki, Finland |
| Venla Kurra | Pirkanmaa Hospital District, Tampere, Finland |
| Oskari Heikinheimo | Hospital District of Helsinki and Uusimaa, Helsinki, Finland |
| Ilkka Kalliala | Hospital District of Helsinki and Uusimaa, Helsinki, Finland |
| Laura Kotaniemi-Talonen | Pirkanmaa Hospital District, Tampere, Finland |
| Kari Nieminen | Pirkanmaa Hospital District, Tampere, Finland |
| Päivi Polo | Hospital District of Southwest Finland, Turku, Finland |
| Kaarin Mäkikallio | Hospital District of Southwest Finland, Turku, Finland |
| Eeva Ekholm | Hospital District of Southwest Finland, Turku, Finland |
| Marja Vääräsmäki | Northern Ostrobothnia Hospital District, Oulu, Finland |
| Outi Uimari | Northern Ostrobothnia Hospital District, Oulu, Finland |
| Laure Morin-Papunen | Northern Ostrobothnia Hospital District, Oulu, Finland |
| Marjo Tuppurainen | Northern Savo Hospital District, Kuopio, Finland |
| Katja Kivinen | Institute for Molecular Medicine Finland, HiLIFE, University of Helsinki, Finland |
| Elisabeth Widen | Institute for Molecular Medicine Finland, HiLIFE, University of Helsinki, Finland |
| Taru Tukiainen | Institute for Molecular Medicine Finland, HiLIFE, University of Helsinki, Finland |
| Mary Pat Reeve | Institute for Molecular Medicine Finland, HiLIFE, University of Helsinki, Finland |
| Mark Daly | Institute for Molecular Medicine Finland, HiLIFE, University of Helsinki, Finland |
| Liu Aoxing | Institute for Molecular Medicine Finland, HiLIFE, University of Helsinki, Finland |
| Eija Laakkonen | University of Jyväskylä, Jyväskylä, Finland |
| Niko Välimäki | University of Helsinki, Helsinki, Finland |
| Lauri Aaltonen | Hospital District of Helsinki and Uusimaa, Helsinki, Finland |
| Johannes Kettunen | Northern Ostrobothnia Hospital District, Oulu, Finland |
| Mikko Arvas | Finnish Red Cross Blood Service, Helsinki, Finland |
| Jeffrey Waring | Abbvie, Chicago, IL, United States |
| Bridget Riley-Gillis | Abbvie, Chicago, IL, United States |

| | |
|---|---|
| Mengzhen Liu | Abbvie, Chicago, IL, United States |
| Janet Kumar | GlaxoSmithKline, Brentford, United Kingdom |
| Kirsi Auro | GlaxoSmithKline, Brentford, United Kingdom |
| Andrea Ganna | Institute for Molecular Medicine Finland, HiLIFE, University of Helsinki, Finland |
| Sarah Pendergrass | Genentech, San Francisco, CA, United States |

##### FinnGen Analysis working group

| | |
|---|---|
| Justin Wade Davis | Abbvie, Chicago, IL, United States |
| Bridget Riley-Gillis | Abbvie, Chicago, IL, United States |
| Danjuma Quarless | Abbvie, Chicago, IL, United States |
| Fedik Rahimov | Abbvie, Chicago, IL, United States |
| Sahar Esmaeeli | Abbvie, Chicago, IL, United States |
| Slavé Petrovski | Astra Zeneca, Cambridge, United Kingdom |
| Eleonor Wigmore | Astra Zeneca, Cambridge, United Kingdom |
| Adele Mitchell | Biogen, Cambridge, MA, United States |
| Benjamin Sun | Biogen, Cambridge, MA, United States |
| Ellen Tsai | Biogen, Cambridge, MA, United States |
| Denis Baird | Biogen, Cambridge, MA, United States |
| Paola Bronson | Biogen, Cambridge, MA, United States |
| Ruoyu Tian | Biogen, Cambridge, MA, United States |
| Stephanie Loomis | Biogen, Cambridge, MA, United States |
| Yunfeng Huang | Biogen, Cambridge, MA, United States |
| Joseph Maranville | Celgene, Summit, NJ, United States/ Bristol Myers Squibb, New York, NY, United States |
| Shameek Biswas | Celgene, Summit, NJ, United States/ Bristol Myers Squibb, New York, NY, United States |
| Elmutaz Mohammed | Celgene, Summit, NJ, United States/ Bristol Myers Squibb, New York, NY, United States |
| Samir Wadhawan | Celgene, Summit, NJ, United States/ Bristol Myers Squibb, New York, NY, United States |
| Erika Kvikstad | Celgene, Summit, NJ, United States/ Bristol Myers Squibb, New York, NY, United States |
| Minal Caliskan | Celgene, Summit, NJ, United States/ Bristol Myers Squibb, New York, NY, United States |
| Diana Chang | Genentech, San Francisco, CA, United States |
| Julie Hunkapiller | Genentech, San Francisco, CA, United States |
| Tushar Bhangale | Genentech, San Francisco, CA, United States |

Natalie Bowers Genentech, San Francisco, CA, United States

Sarah Pendergrass Genentech, San Francisco, CA, United States

Kirill Shkura Merck, Kenilworth, NJ, United States

Victor Neduva Merck, Kenilworth, NJ, United States

Xing Chen Pfizer, New York, NY, United States

Åsa Hedman Pfizer, New York, NY, United States

Karen S King GlaxoSmithKline, Brentford, United Kingdom

Padhraig Gormley GlaxoSmithKline, Brentford, United Kingdom

Jimmy Liu GlaxoSmithKline, Brentford, United Kingdom

Clarence Wang Sanofi, Paris, France

Ethan Xu Sanofi, Paris, France

Franck Auge Sanofi, Paris, France

Clement Chatelain Sanofi, Paris, France

Deepak Rajpal Sanofi, Paris, France

Dongyu Liu Sanofi, Paris, France

Katherine Call Sanofi, Paris, France

Tai-He Xia Sanofi, Paris, France

Beryl Cummings Maze Therapeutics, San Francisco, CA, United States

Matt Brauer Maze Therapeutics, San Francisco, CA, United States

Huilei Xu Novartis, Basel, Switzerland

Amy Cole Novartis, Basel, Switzerland

Jonathan Chung Novartis, Basel, Switzerland

Jaison Jacob Novartis, Basel, Switzerland

Katrina de Lange Novartis, Basel, Switzerland

Jonas Zierer Novartis, Basel, Switzerland

Mitja Kurki Institute for Molecular Medicine Finland, HiLIFE, University of Helsinki, Finland / Broad Institute, Cambridge, MA, United States

Samuli Ripatti Institute for Molecular Medicine Finland, HiLIFE, University of Helsinki, Finland

Mark Daly Institute for Molecular Medicine Finland, HiLIFE, University of Helsinki, Finland

Juha Karjalainen Institute for Molecular Medicine Finland, HiLIFE, University of Helsinki, Finland / Broad Institute, Cambridge, MA, United States
Aki Havulinna Institute for Molecular Medicine Finland, HiLIFE, University of Helsinki, Finland
Juha Mehtonen Institute for Molecular Medicine Finland, HiLIFE, University of Helsinki, Finland
Priit Palta Institute for Molecular Medicine Finland, HiLIFE, University of Helsinki, Finland
Shabbeer Hassan Institute for Molecular Medicine Finland, HiLIFE, University of Helsinki, Finland
Pietro Della Briotta Parolo Institute for Molecular Medicine Finland, HiLIFE, University of Helsinki, Finland
Wei Zhou Broad Institute, Cambridge, MA, United States
Mutaamba Maasha Broad Institute, Cambridge, MA, United States
Shabbeer Hassan Institute for Molecular Medicine Finland, HiLIFE, University of Helsinki, Finland
Susanna Lemmelä Institute for Molecular Medicine Finland, HiLIFE, University of Helsinki, Finland
Manuel Rivas University of Stanford, Stanford, CA, United States
Aarno Palotie Institute for Molecular Medicine Finland, HiLIFE, University of Helsinki, Finland
Arto Lehisto Institute for Molecular Medicine Finland, HiLIFE, University of Helsinki, Finland
Andrea Ganna Institute for Molecular Medicine Finland, HiLIFE, University of Helsinki, Finland
Vincent Llorens Institute for Molecular Medicine Finland, HiLIFE, University of Helsinki, Finland
Hannele Laivuori Institute for Molecular Medicine Finland, HiLIFE, University of Helsinki, Finland
Mari E Niemi Institute for Molecular Medicine Finland, HiLIFE, University of Helsinki, Finland
Taru Tukiainen Institute for Molecular Medicine Finland, HiLIFE, University of Helsinki, Finland
Mary Pat Reeve Institute for Molecular Medicine Finland, HiLIFE, University of Helsinki, Finland
Henrike Heyne Institute for Molecular Medicine Finland, HiLIFE, University of Helsinki, Finland
Nina Mars Institute for Molecular Medicine Finland, HiLIFE, University of Helsinki, Finland
Kimmo Palin University of Helsinki, Helsinki, Finland
Javier Garcia-Tabuenca University of Tampere, Tampere, Finland
Harri Siirtola University of Tampere, Tampere, Finland
Tuomo Kiiskinen Institute for Molecular Medicine Finland, HiLIFE, University of Helsinki, Finland
Jiwoo Lee Institute for Molecular Medicine Finland, HiLIFE, University of Helsinki, Finland / Broad Institute, Cambridge, MA, United States
Kristin Tsuo Institute for Molecular Medicine Finland, HiLIFE, University of Helsinki, Finland / Broad Institute, Cambridge, MA, United States

Amanda Elliott Institute for Molecular Medicine Finland, HiLIFE, University of Helsinki, Finland / Broad Institute, Cambridge, MA, United States

Kati Kristiansson THL Biobank / The National Institute of Health and Welfare Helsinki, Finland

Mikko Arvas Finnish Red Cross Blood Service / Finnish Hematology Registry and Clinical Biobank, Helsinki, Finland

Kati Hyvärinen Finnish Red Cross Blood Service, Helsinki, Finland

Jarmo Ritari Finnish Red Cross Blood Service, Helsinki, Finland

Miika Koskinen Helsinki Biobank / Helsinki University and Hospital District of Helsinki and Uusimaa, Helsinki

Olli Carpén Helsinki Biobank / Helsinki University and Hospital District of Helsinki and Uusimaa, Helsinki

Johannes Kettunen Northern Finland Biobank Borealis / University of Oulu / Northern Ostrobothnia Hospital District, Oulu, Finland

Katri Pylkäs University of Oulu, Oulu, Finland

Marita Kalaoja University of Oulu, Oulu, Finland

Minna Karjalainen University of Oulu, Oulu, Finland

Tuomo Mantere Northern Finland Biobank Borealis / University of Oulu / Northern Ostrobothnia Hospital District, Oulu, Finland

Eeva Kangasniemi Finnish Clinical Biobank Tampere / University of Tampere / Pirkanmaa Hospital District, Tampere, Finland

Sami Heikkinen University of Eastern Finland, Kuopio, Finland

Arto Mannermaa Biobank of Eastern Finland / University of Eastern Finland / Northern Savo Hospital District, Kuopio, Finland

Eija Laakkonen University of Jyväskylä, Jyväskylä, Finland

Samuel Heron University of Turku, Turku, Finland

Dhanaprakash Jambulingam University of Turku, Turku, Finland

Venkat Subramaniam Rathinakannan University of Turku, Turku, Finland

Nina Pitkänen Auria Biobank / University of Turku / Hospital District of Southwest Finland, Turku, Finland

#### Biobank directors

Lila Kallio Auria Biobank / University of Turku / Hospital District of Southwest Finland, Turku, Finland

Sirpa Soini THL Biobank / The National Institute of Health and Welfare Helsinki, Finland

Jukka Partanen Finnish Red Cross Blood Service / Finnish Hematology Registry and Clinical Biobank, Helsinki, Finland

Eero Punkka Helsinki Biobank / Helsinki University and Hospital District of Helsinki and Uusimaa, Helsinki

Raisa Serpi Northern Finland Biobank Borealis / University of Oulu / Northern Ostrobothnia Hospital District, Oulu, Finland

Johanna Mäkelä Finnish Clinical Biobank Tampere / University of Tampere / Pirkanmaa Hospital District, Tampere, Finland

Veli-Matti Kosma Biobank of Eastern Finland / University of Eastern Finland / Northern Savo Hospital District, Kuopio, Finland

Teijo Kuopio Central Finland Biobank / University of Jyväskylä / Central Finland Health Care District, Jyväskylä, Finland

#### FinnGen Teams

**Administration**

Anu Jalanko Institute for Molecular Medicine Finland, HiLIFE, University of Helsinki, Finland

Huei-Yi Shen Institute for Molecular Medicine Finland, HiLIFE, University of Helsinki, Finland

Risto Kajanne Institute for Molecular Medicine Finland, HiLIFE, University of Helsinki, Finland

Mervi Aavikko Institute for Molecular Medicine Finland, HiLIFE, University of Helsinki, Finland

**Analysis**

Mitja Kurki Institute for Molecular Medicine Finland, HiLIFE, University of Helsinki, Finland / Broad Institute, Cambridge, MA, United States

Juha Karjalainen Institute for Molecular Medicine Finland, HiLIFE, University of Helsinki, Finland / Broad Institute, Cambridge, MA, United States

| | |
|---|---|
| Pietro Della Briotta Parolo | Institute for Molecular Medicine Finland, HiLIFE, University of Helsinki, Finland |
| Arto Lehisto | Institute for Molecular Medicine Finland, HiLIFE, University of Helsinki, Finland |
| Juha Mehtonen | Institute for Molecular Medicine Finland, HiLIFE, University of Helsinki, Finland |
| Wei Zhou | Broad Institute, Cambridge, MA, United States |
| Masahiro Kanai | Broad Institute, Cambridge, MA, United States |
| Mutaamba Maasha | Broad Institute, Cambridge, MA, United States |

**Clinical Endpoint Development**

| | |
|---|---|
| Hannele Laivuori | Institute for Molecular Medicine Finland, HiLIFE, University of Helsinki, Finland |
| Aki Havulinna | Institute for Molecular Medicine Finland, HiLIFE, University of Helsinki, Finland |
| Susanna Lemmelä | Institute for Molecular Medicine Finland, HiLIFE, University of Helsinki, Finland |
| Tuomo Kiiskinen | Institute for Molecular Medicine Finland, HiLIFE, University of Helsinki, Finland |
| L. Elisa Lahtela | Institute for Molecular Medicine Finland, HiLIFE, University of Helsinki, Finland |
| Matti Peura | Institute for Molecular Medicine Finland, HiLIFE, University of Helsinki, Finland |

**Communication**

| | |
|---|---|
| Mari Kaunisto | Institute for Molecular Medicine Finland, HiLIFE, University of Helsinki, Finland |

**Data Management and IT Infrastructure**

| | |
|---|---|
| Elina Kilpeläinen | Institute for Molecular Medicine Finland, HiLIFE, University of Helsinki, Finland |
| Timo P. Sipilä | Institute for Molecular Medicine Finland, HiLIFE, University of Helsinki, Finland |
| Georg Brein | Institute for Molecular Medicine Finland, HiLIFE, University of Helsinki, Finland |
| Oluwaseun A. Dada | Institute for Molecular Medicine Finland, HiLIFE, University of Helsinki, Finland |
| Awaisa Ghazal | Institute for Molecular Medicine Finland, HiLIFE, University of Helsinki, Finland |
| Anastasia Shcherban | Institute for Molecular Medicine Finland, HiLIFE, University of Helsinki, Finland |

**Genotyping**

| | |
|---|---|
| Kati Donner | Institute for Molecular Medicine Finland, HiLIFE, University of Helsinki, Finland |
| Timo P. Sipilä | Institute for Molecular Medicine Finland, HiLIFE, University of Helsinki, Finland |

**Sample Collection Coordination**

Anu Loukola	Helsinki Biobank / Helsinki University and Hospital District of Helsinki and Uusimaa, Helsinki

**Sample Logistics**

Päivi Laiho	THL Biobank / The National Institute of Health and Welfare Helsinki, Finland

Tuuli Sistonen	THL Biobank / The National Institute of Health and Welfare Helsinki, Finland

Essi Kaiharju	THL Biobank / The National Institute of Health and Welfare Helsinki, Finland

Markku Laukkanen	THL Biobank / The National Institute of Health and Welfare Helsinki, Finland

Elina Järvensivu	THL Biobank / The National Institute of Health and Welfare Helsinki, Finland

Sini Lähteenmäki	THL Biobank / The National Institute of Health and Welfare Helsinki, Finland

Lotta Männikkö	THL Biobank / The National Institute of Health and Welfare Helsinki, Finland

Regis Wong	THL Biobank / The National Institute of Health and Welfare Helsinki, Finland

**Registry Data Operations**

Hannele Mattsson	THL Biobank / The National Institute of Health and Welfare Helsinki, Finland

Kati Kristiansson	THL Biobank / The National Institute of Health and Welfare Helsinki, Finland

Susanna Lemmelä	Institute for Molecular Medicine Finland, HiLIFE, University of Helsinki, Finland

Sami Koskelainen	THL Biobank / The National Institute of Health and Welfare Helsinki, Finland

Tero Hiekkalinna	THL Biobank / The National Institute of Health and Welfare Helsinki, Finland

Teemu Paajanen	THL Biobank / The National Institute of Health and Welfare Helsinki, Finland

**Sequencing Informatics**

Priit Palta	Institute for Molecular Medicine Finland, HiLIFE, University of Helsinki, Finland

Kalle Pärn	Institute for Molecular Medicine Finland, HiLIFE, University of Helsinki, Finland

Shuang Luo	Institute for Molecular Medicine Finland, HiLIFE, University of Helsinki, Finland

Vishal Sinha	Institute for Molecular Medicine Finland, HiLIFE, University of Helsinki, Finland

**Trajectory Team**

Tarja Laitinen Pirkanmaa Hospital District, Tampere, Finland

Harri Siirtola University of Tampere, Tampere, Finland

Javier Gracia-Tabuenca University of Tampere, Tampere, Finland

**Data protection officer**

Tero Jyrhämä Institute for Molecular Medicine Finland, HiLIFE, University of Helsinki, Finland

**FinBB - Finnish biobank cooperative**

Marco Hautalahti

Laura Mustaniemi

Mirkka Koivusalo

Sarah Smith

Tom Southerington
