## Supplementary Fig 1 for "Leveraging Northern European population history; novel low frequency variants for polycystic ovary syndrome"

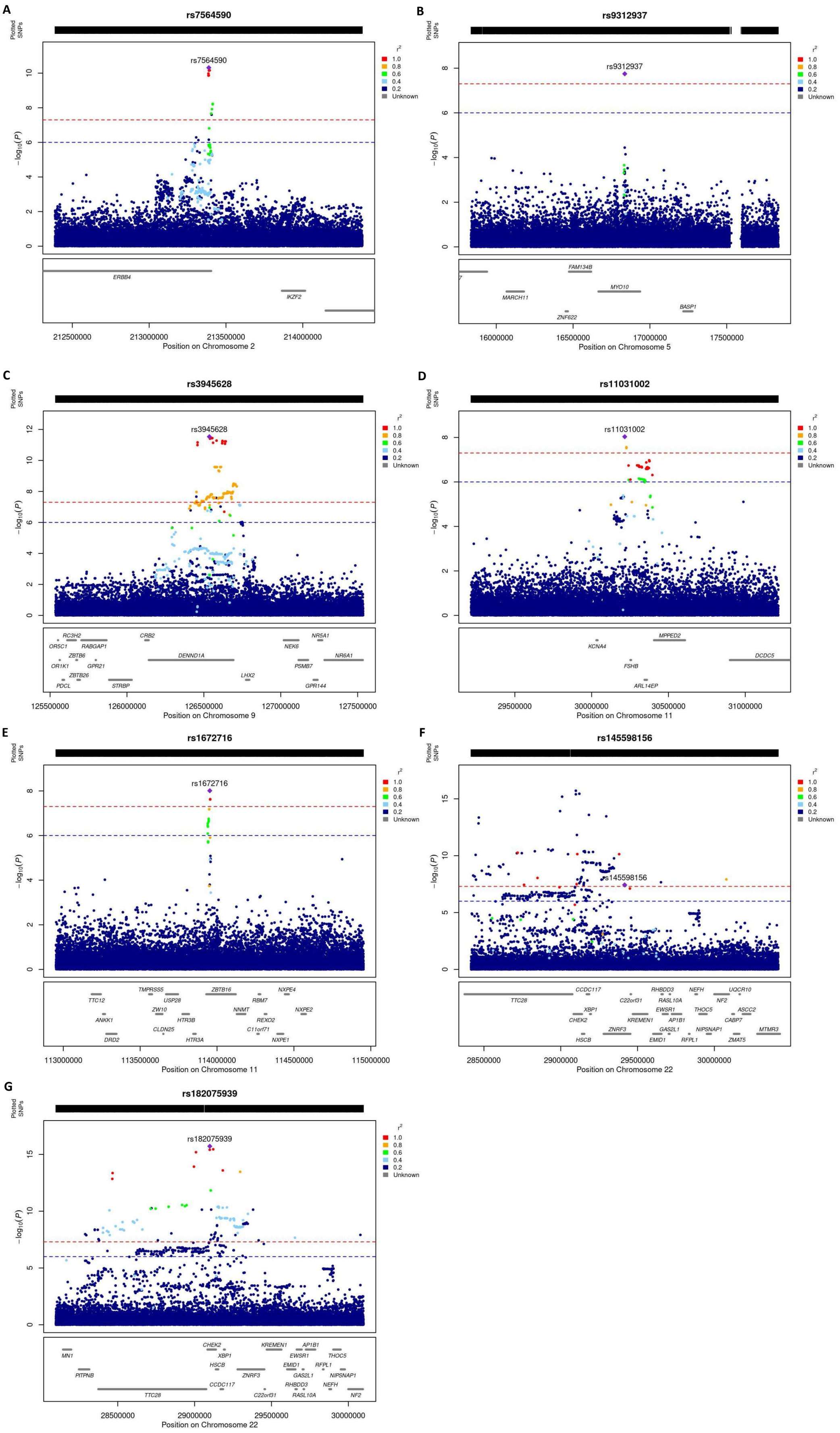


**Supplementary Fig 1**. **Regional association plots of genome-wide significant variants.** Regional plots of association (left y-axis) and recombination rates (right y-axis) for the chromosomes (a) 2q34, (b) 5p15, (c) 9q33, (d) 11.p14, (e) 11q23, (f) 22q12, and (g) 22q12 loci after meta-analyses. The lead SNP in each locus is labeled and marked in purple. All other SNPs are color coded according to the strength of LD with the top SNP (as measured by $r^2$ in the SiSu v3 reference panel).
