## Supplementary Fig 2 for "Leveraging Northern European population history; novel low frequency variants for polycystic ovary syndrome"

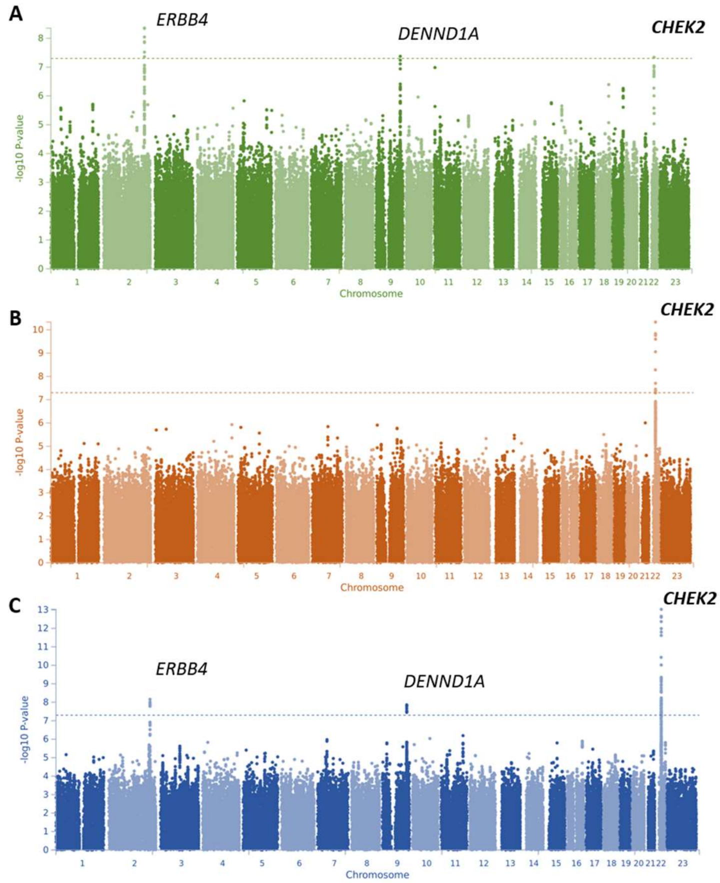


**Supplementary Fig 2. Manhattan plot for age- and BMI-adjusted GWAS in the Finnish dataset (A) the Estonian dataset (B) and the joint GWAS meta-analysis of PCOS (C).** The novel candidates are highlighted in bold. The y axis represents -log(two-sided P values) for association between variants and PCOS from meta-analysis using an inverse-variance weighted fixed effects model. The horizontal dashed line represents the threshold for genome-wide significance.
